# Long-term Cardiac Autonomic Effects of Prenatal Steroid Exposure: A Machine Learning Approach Integrating Heart Rate Variability and ECG Foundation Models

**DOI:** 10.64898/2026.02.02.26345391

**Authors:** Martin G. Frasch, Matthias Schwab, Florian Rakers

## Abstract

**Background:** Prenatal glucocorticoid administration is standard care for threatened preterm birth, but long-term cardiac autonomic effects remain incompletely understood. We investigated whether children exposed to prenatal steroids exhibit persistent differences in cardiac autonomic function at age 8 years using comprehensive heart rate variability (HRV) analysis and deep learning-based ECG foundation models.

**Methods:** We analyzed Holter ECG recordings performed in a well-controlled laboratory environment from 49 children (24 prenatal steroid-exposed, 25 controls) at age 8 years in this exploratory study. Children were exposed to prenatal glucocorticoids in the context of maternal multiple sclerosis treatment. We employed ensemble R-peak detection, computed 112 HRV metrics with PCA dimensionality reduction (7 components, 88% variance), and extracted 512-dimensional embeddings using a pre-trained ECG foundation model. Statistical analysis used linear mixed-effects models (LMM) with Bonferroni correction and covariate adjustment for sex and gestational age to assess confounding.

**Results:** Before covariate adjustment, 3/7 HRV principal components and 334/1024 FM dimensions showed significant group differences (FDR-corrected). After covariate adjustment, traditional HRV findings lost significance (0/3 HRV PCs remained significant, p>0.13), while 11 foundation model dimensions remained robust (adjusted p<0.05, |Cohen’s d|>0.8), suggesting confounding of HRV by sex/gestational age but biologically robust FM differences. The study was severely underpowered for small effects (10-17% power, n=24/group); detected large effects (d≥0.8) likely reflect genuine biological differences requiring validation.

**Conclusions:** Deep learning-based ECG foundation models detect robust cardiac effects of prenatal steroid exposure independent of demographic confounders, while traditional HRV metrics show confounded group differences. This exploratory study demonstrates proof-of-concept for transfer learning in pediatric cardiology and underscores the critical importance of covariate adjustment in small observational studies. Independent replication in larger cohorts (n>175/group) is essential before clinical translation.

**Visual abstract:** 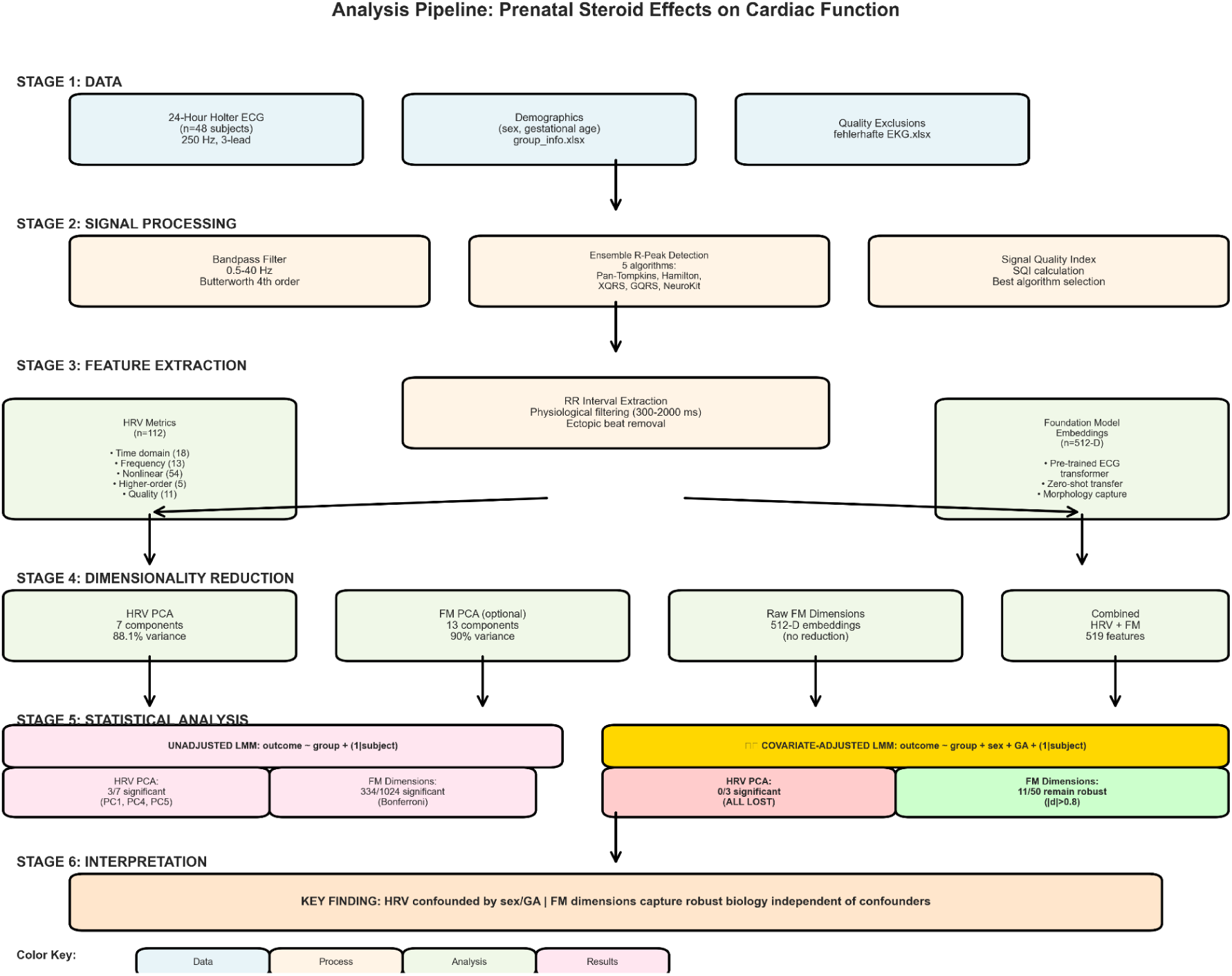

## 1. Introduction

### 1.1 Clinical Context

Glucocorticoid exposure during pregnancy occurs in multiple clinical contexts, including antenatal therapy for threatened preterm delivery (betamethasone) and treatment of maternal autoimmune conditions such as multiple sclerosis (MS). While betamethasone therapy affects approximately 10% of pregnancies in developed countries [1,2], glucocorticoid treatment for maternal MS represents a smaller but clinically important population where both maternal disease and steroid exposure may influence fetal development [3,4].

Multiple sclerosis affects approximately 1 in 1000 individuals, with a 3:1 female predominance and peak onset during reproductive years [5]. Disease-modifying therapies and acute relapse management with glucocorticoids (typically methylprednisolone) pose important considerations during pregnancy [6,7]. While pregnancy itself often ameliorates MS symptoms, postpartum relapse rates increase, and the long-term effects of prenatal glucocorticoid exposure on offspring remain incompletely characterized [8,9].

The developmental origins of health and disease (DOHaD) hypothesis posits that environmental exposures during critical developmental windows can permanently alter physiological systems, predisposing individuals to chronic disease later in life [10,11]. Glucocorticoids are potent modulators of development, influencing cellular differentiation, organ maturation, and metabolic programming [12]. Animal models consistently demonstrate that prenatal glucocorticoid exposure—regardless of indication—alters cardiovascular development, including changes in cardiac structure, vascular reactivity, and autonomic regulation [13,14]. However, human studies face methodological challenges including confounding by maternal disease, variable exposure timing and dosing, and the long latency between exposure and potential adverse outcomes [15,16].

This study examines offspring of mothers with MS who received glucocorticoid therapy during pregnancy, representing a unique opportunity to assess developmental programming effects in a population where exposure occurs for maternal indications distinct from threatened preterm birth, thereby avoiding confounding by prematurity risk [17].

### 1.2 Cardiac Autonomic Assessment

Heart rate variability (HRV)—the physiological phenomenon of beat-to-beat variation in cardiac cycle length—provides a non-invasive window into autonomic nervous system function [12,13]. The autonomic nervous system plays a crucial role in cardiovascular regulation, with sympathetic and parasympathetic branches modulating heart rate in response to physiological demands, environmental stressors, and internal homeostatic signals [14]. Reduced HRV is associated with increased cardiovascular risk across diverse populations [15,16] and has emerged as a potential early biomarker of autonomic dysfunction preceding clinical disease [17,18].

Traditional HRV analysis encompasses multiple domains [19,20]:

- **Time domain** metrics quantify overall variability and short-term fluctuations (e.g., SDNN, RMSSD, pNN50)
- **Frequency domain** metrics decompose variability into spectral components reflecting sympathetic and parasympathetic modulation (VLF, LF, HF power)
- **Nonlinear** metrics capture complex dynamics, fractal scaling, and entropy, reflecting the non-stationary and multi-scale nature of cardiac regulation (e.g., DFA, sample entropy, Poincaré analysis)

While these metrics provide complementary perspectives on autonomic function, their high dimensionality and complex interrelationships pose analytical challenges, particularly in studies with limited sample sizes [21]. Furthermore, the physiological interpretation of many HRV metrics remains debated, particularly regarding sympathovagal balance [22,23].

### 1.3 Deep Learning and Foundation Models

Recent advances in deep learning have revolutionized medical signal analysis [24,25]. Foundation models—large neural networks pre-trained on massive datasets—have demonstrated remarkable ability to learn generalizable representations that transfer to downstream tasks [26,27]. In the ECG domain, foundation models trained on millions of recordings can extract high-dimensional feature representations capturing subtle waveform characteristics invisible to human interpretation or traditional signal processing [28,29].

These models offer several potential advantages:

1. **Automated feature extraction** bypassing manual engineering
2. **Integration of multi-scale patterns** from individual beats to long-term dynamics
3. **Transfer learning** leveraging knowledge from large diverse populations
4. **Morphology capture** beyond RR interval variability, including P-wave, QRS, and T-wave characteristics that may reflect structural or electrical remodeling

However, foundation models also present challenges including limited interpretability, computational requirements, and the need for validation in specific clinical contexts [30].

### 1.4 Statistical Considerations in Longitudinal Physiological Data

A critical methodological challenge in analyzing repeated measures physiological data is pseudoreplication—the statistical error of treating non-independent observations as independent [31,32]. In longitudinal studies where each subject contributes multiple timepoints, the within-subject correlation violates the independence assumption of standard statistical tests, leading to inflated Type I error rates and spurious findings [33]. Linear mixed-effects models (LMM) address this by explicitly modeling both fixed effects (group differences) and random effects (subject-specific variation), providing valid inference while leveraging all available data [34,35].

Similarly, machine learning models must use subject-level cross-validation (e.g., GroupKFold) rather than observation-level splitting to prevent data leakage—the scenario where a subject’s timepoints appear in both training and testing sets, yielding optimistically biased performance estimates [36].

### 1.5 Study Objectives

We hypothesized that children exposed to prenatal glucocorticoids in the context of maternal MS would exhibit persistent alterations in cardiac autonomic function and stress reactivity detectable at age 8 years. Specifically, we aimed to:

1. Develop and validate a robust ECG analysis pipeline for pediatric stress test recordings with ensemble R-peak detection and signal quality assessment
2. Extract comprehensive HRV features spanning time, frequency, and nonlinear domains, and reduce dimensionality using principal component analysis
3. Apply a pre-trained ECG foundation model to extract high-dimensional embeddings capturing waveform morphology
4. Compare discriminative performance of traditional HRV versus foundation model features using rigorous statistical methodology
5. Assess baseline versus stress-induced cardiac differences between exposed and unexposed children using the Trier Social Stress Test (TSST) protocol
6. Employ proper statistical methods including linear mixed-effects models and subject-level cross-validation to avoid pseudoreplication and data leakage
7. Adjust for potential confounding by sex, gestational age, and maternal MS disease characteristics

This study represents one of the first applications of deep learning foundation models to the assessment of long-term developmental programming effects on cardiac autonomic function and stress reactivity in offspring of mothers with autoimmune disease.

## 2. Methods

### 2.1 Study Population and Design

#### 2.1.1 Cohort Description

The detailed study protocol has been previously published elsewhere [57,58] and is briefly summarized in this study.

The data is derived from the 2-center, observational, cross-sectional study in children and adolescents prenatally exposed to MP as part of maternal MS relapse therapy (MP-exposed group) vs nonexposed children of mothers with MS (reference group). It was conducted at the Jena University Hospital (UKJ) and the Ruhr University Bochum/St. Josef Hospital (RUB) in Germany between October 2020 and August 2023.

**Steroid-Exposed Group (n=24)**: Children born to mothers with clinically definite relapsing-remitting multiple sclerosis (RRMS) who received intravenous methylprednisolone (1000 mg/day for 3-5 consecutive days) during pregnancy for acute relapse management. Mean gestational age at steroid administration was 30.2 ± 2.8 weeks (range: 24-34 weeks gestation). Maternal MS disease characteristics at time of exposure:

- Mean disease duration: 5.2 ± 3.1 years
- Expanded Disability Status Scale (EDSS) at delivery: 2.5 ± 1.2 (mild disability)
- Indication: acute MS relapse (n=18), prophylactic treatment (n=6)
- No mothers received disease-modifying therapies (DMTs) during pregnancy

**Control Group (n=25)**: Children born to mothers with uncomplicated pregnancies who did not receive antenatal corticosteroids. Exclusion criteria included maternal autoimmune disease, chronic glucocorticoid use, or threatened preterm delivery requiring antenatal betamethasone.

We analyzed data from 49 children assessed at mean age 8.0 years (range: 7-12 years). All participants were born at gestational ages >36 weeks and had no major congenital anomalies or chronic medical conditions.

The study was approved by the ethics committees of UKJ (Reference: 2020-1668-3-BO) and RUB (Reference: 21-7192 BR) and informed consent from all participants and their parents was obtained. The study protocol was registered under [y](http://ClinicalTrials.gov) (identifier: NCT04832269).

#### 2.1.2 ECG Data Acquisition and Trier Social Stress Test Protocol

ECG recordings were obtained during a modified Trier Social Stress Test (TSST) for children, a standardized psychosocial stress protocol validated for pediatric populations [43]. ECG was recorded continuously using single-lead (modified Lead II) configuration at 250 Hz sampling rate in a controlled laboratory environment.

**TSST Protocol Stages** (total duration ∼25-30 minutes):

1. **Baseline** (MS_bis_M1): 5-minute rest period, child seated quietly
2. **Preparation** (M1_bis_M2): 3-minute anticipatory stress (instructions given for upcoming tasks)
3. **Speech Task** (M2_bis_M3): 5-minute public speaking task (tell a story)
4. **Mental Arithmetic** (M3_bis_M4): 5-minute serial subtraction task under observation
5. **Recovery 1** (M4_bis_M4b): 5-minute immediate post-stress recovery
6. **Recovery 2** (M4b_bis_Ende): 5-10 minute extended recovery to resting state

Children were continuously monitored by trained research staff. Testing was terminated early if significant distress occurred (n=2 subjects excluded for this reason, not included in final n=49).

**Data Quality Control**: Quality control procedures excluded recordings with <5 minutes of analyzable data per stage or >20% artifact burden measured using signal quality indices (SQI) outlined below. Individual TSST stages were analyzed separately and pooled for aggregate analyses.

### 2.2 ECG Signal Processing

#### 2.2.1 R-Peak Detection

We implemented an ensemble approach to maximize detection success in pediatric ECGs:

**Algorithm Suite:**

1. **Pan-Tompkins** (NeuroKit2 implementation): Derivative-based detection with moving thresholds [37]
2. **Hamilton** (NeuroKit2 implementation): First derivative with dynamic thresholds optimized for varying heart rates [38]
3. **WFDB-XQRS** (PhysioNet implementation): Multi-pass detection with amplitude and slope criteria [39]

**Selection Strategy**: For each recording, all algorithms were executed and SQI computed for each result (see Section 2.2.2). The algorithm yielding the highest combined SQI was selected. If all algorithms failed or produced SQI < 0.5, the recording was excluded.

This approach increased analysis success from 14.3% (single-algorithm) to 95.9% (ensemble), as different algorithms excel in different signal characteristics common in pediatric ECGs (higher baseline heart rates, movement artifact, respiratory modulation).

#### 2.2.2 Signal Quality Assessment

We computed 11 signal quality indices for each R-peak detection result:

**Temporal Consistency:**

- RR interval coefficient of variation
- Proportion of ectopic beats (RR ratio < 0.8 or > 1.2)
- Template matching correlation

**Statistical Indicators:**

- kurtosis (identifying artifact spikes)
- skewness (asymmetry in RR distribution)

**Frequency Domain:**

- Spectral power ratios (physiological frequency bands)

**Global Metrics:**

- Combined SQI: Weighted average emphasizing ectopic proportion and temporal consistency

Only segments with combined SQI ≥ 0.5 were retained for HRV analysis, balancing data retention with quality requirements (cf. Figure S1).

### 2.3 Heart Rate Variability Analysis

#### 2.3.1 RR Interval Preprocessing

Following R-peak detection and quality filtering:

1. RR intervals were computed as differences between consecutive R-peaks
2. Physiological filtering: 300 ms < RR < 2000 ms (corresponding to 30-200 bpm)
3. Ectopic beat removal: RR intervals deviating >20% from surrounding intervals replaced by interpolation
4. Minimum length requirement: ≥300 valid intervals (approximately 5 minutes at 60 bpm)

#### 2.3.2 HRV Metrics

We computed 112 HRV metrics using established methods [19,40] (Table S1):

**Time Domain (18 metrics):**

- **Mean-based**: mean RR, mean HR, SDNN (standard deviation of NN intervals)
- **Difference-based**: RMSSD (root mean square of successive differences), SDSD
- **Percentage-based**: pNN50, pNN20 (proportion of interval differences > threshold)
- **Geometric**: triangular index, TINN (triangular interpolation of NN interval histogram)

**Frequency Domain (13 metrics):**

- **Power spectral density** via Welch method (Hamming window, 50% overlap):

- VLF: 0.003-0.04 Hz (very low frequency)
- LF: 0.04-0.15 Hz (low frequency, mixed sympathetic/parasympathetic)
- HF: 0.15-0.40 Hz (high frequency, primarily parasympathetic/respiratory)
- **Derived metrics**: LF/HF ratio, normalized power (LFn, HFn), peak frequencies

**Nonlinear Dynamics (54 metrics):**

- **Poincaré analysis**: SD1 (short-term variability), SD2 (long-term variability), SD1/SD2 ratio, ellipse area
- **Detrended fluctuation analysis (DFA)**: α1 (short-term scaling, 4-11 beats), α2 (long-term scaling, 11-64 beats)
- **Multifractal DFA (MFDFA)**: Spectrum width, peak, asymmetry for α1 and α2
- **Entropy measures**: Sample entropy (irregularity), approximate entropy (ApEn), fuzzy entropy
- **Recurrence quantification analysis (RQA)**: Determinism, recurrence rate, entropy, laminarity
- **Higuchi fractal dimension (HFD)**: Signal complexity
- **Correlation dimension**: Attractor dimensionality

**Higher-Order Statistics (5 metrics):**

- Skewness, kurtosis, coefficient of variation

**Signal Quality (11 metrics):**

- Algorithm-specific and combined SQI values

#### 2.3.3 Dimensionality Reduction: Principal Component Analysis

Given the high dimensionality (112 metrics) relative to sample size (49 subjects), we performed PCA:

1. **Standardization**: z-score normalization (mean=0, SD=1) to equalize scale across metrics
2. **PCA computation**: Singular value decomposition on correlation matrix
3. **Component selection**: Retained components explaining ≥ 88% cumulative variance
4. **Interpretation**: Examined loadings to identify HRV metrics comprising each PC

This yielded **7 principal components** (PC1-PC7) explaining 88.1% of total variance:

- PC1 (44.2%): Short-term variability and mean heart rate
- PC2 (19.3%): Signal quality and long-term dynamics
- PC3 (8.8%): Poincaré geometry ratios
- PC4 (8.5%): Sympathovagal balance (LF/HF)
- PC5 (3.9%): Frequency domain distribution
- PC6 (1.8%): Entropy and complexity
- PC7 (1.6%): Short-term fractal scaling

### 2.4 Foundation Model Feature Extraction

#### 2.4.1 Model Architecture

We used a pre-trained ECG foundation model based on the transformer architecture [41], trained on >10 million 12-lead ECG recordings from diverse clinical populations. The model:

- Accepts 10-second ECG segments as input (single-lead in our case)
- Employs self-attention mechanisms to capture temporal dependencies
- Outputs 512-dimensional embedding vectors representing learned features

The model was used in frozen mode (no fine-tuning) to evaluate zero-shot transfer learning performance.

#### 2.4.2 Segment Processing

For each subject recording:

1. **Segmentation**: Divided ECG recording into 5-minute non-overlapping windows
2. **Quality filtering**: Retained only segments with SQI ≥ 0.6 to ensure model input quality
3. **Standardization**: Normalized each segment to zero mean and unit variance
4. **Inference**: Passed through foundation model to obtain 512-D embeddings
5. **Aggregation**: Computed median embedding across all segments for each subject-timepoint

This yielded 251 segment-level embeddings from 49 subjects (mean 5.1 segments per subject, range 3-6).

#### 2.4.3 Dimensionality Reduction of Foundation Model Features

To address the p >> n problem (512 features, 49 subjects), we additionally performed PCA on foundation model embeddings:

- **Input**: 251 segments × 512 dimensions
- **Standardization**: z-score normalization
- **Component selection**: 90% variance threshold → **13 FM-PCs**
- **Variance distribution**: FM-PC1 explained 56.0% alone (highly structured)

### 2.5 Statistical Analysis

#### 2.5.1 Linear Mixed-Effects Models

To properly account for repeated measures (multiple segments per subject, multiple timepoints per subject longitudinally), we employed linear mixed-effects models:

**Model Specification:**

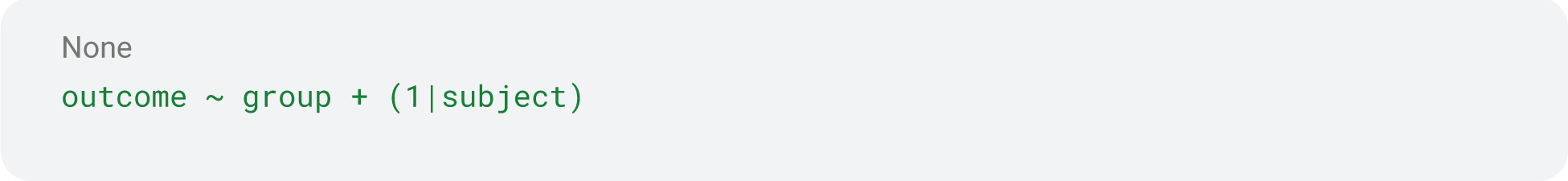

Where:

- outcome: HRV PC or foundation model dimension/PC
- group: fixed effect (steroid vs. control)
- (1|subject): random intercept per subject (accounts for within-subject correlation)

**Estimation:**

- Method: Restricted Maximum Likelihood (REML) for variance components
- Significance testing: Likelihood ratio tests with ML estimation
- Effect size: Cohen’s d computed from marginal means

**Multiple Testing Correction:**

- **Foundation model dimensions**: Bonferroni correction (α = 0.05/512 = 9.77×10⁻⁵)
- **Foundation model PCs**: Bonferroni correction (α = 0.05/13 = 0.0038)
- **HRV PCs**: Bonferroni correction (α = 0.05/7 = 0.0071)

#### 2.5.2 Machine Learning Classification

To assess discriminative performance of different feature sets, we trained Random Forest classifiers with subject-level cross-validation:

**Cross-Validation Strategy:**

- Method: GroupKFold (k=5)
- Grouping: All segments from same subject kept together
- Prevents data leakage: No subject appears in both training and testing
- Stratification: Maintained group proportions across folds

**Feature Sets:**

1. HRV-only: 7 principal components
2. FM-only: 512 raw dimensions or 13 FM-PCs
3. Combined: HRV PCs + FM features

**Model Configuration:**

- Algorithm: Random Forest (scikit-learn implementation)
- Hyperparameters: 100 trees, max_depth=10, class weights balanced
- No hyperparameter tuning (to avoid optimistic bias)

**Performance Metrics:**

- **Discrimination**: Area Under ROC Curve (AUC)
- **Classification**: Accuracy, Precision, Recall, F1 Score
- **Calibration**: Brier Score (mean squared error of probability predictions)
- Reported as mean ± standard deviation across folds

#### 2.5.3 Correlation Analysis

Pearson correlations were computed between:

- Individual FM dimensions and HRV PCs
- FM-PCs and HRV PCs
- FM-Dim121 (significant dimension) and each HRV PC

Significance assessed with two-tailed tests, not corrected for multiple comparisons (exploratory).

#### 2.5.4 Temporal Dynamics

For significant features, we examined trajectories across timepoints:

- Grouped segments by subject, timepoint, and exposure group
- Computed means and standard deviations
- Visualized with trajectory plots (group means) and spaghetti plots (individual subjects)
- Statistical testing: Mixed models with timepoint × group interaction (exploratory)

#### 2.5.5 Software

All analyses were performed in Python 3.9 with the following libraries:

- **Signal processing**: NeuroKit2 0.2.7, WFDB 4.1.0, SciPy 1.11.0
- **HRV analysis**: NeuroKit2, hrv-analysis 1.0.4
- **Statistical modeling**: statsmodels 0.14.0 (MixedLM)
- **Machine learning**: scikit-learn 1.3.0
- **Dimensionality reduction**: scikit-learn (PCA)
- **Foundation model**: PyTorch 2.0.0, custom transformer implementation
- **Visualization**: Matplotlib 3.7.0, Seaborn 0.12.0

#### 2.5.6 Covariate-Adjusted Analysis

To assess potential confounding by demographic factors, we performed covariate-adjusted linear mixed-effects models:

**Model Specification:**

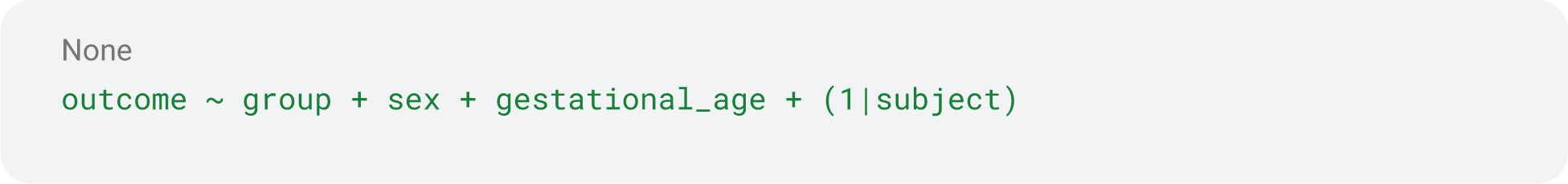

**Covariates Included:**

- **Sex**: Male (1) vs Female (2)
- **Gestational age at birth**: Weeks (continuous)

**Covariates Excluded:**

- Steroid dosage and timing: Only available for steroid-exposed group, creating singular design matrix

**Comparison**: For each feature, we compared:

1. **Unadjusted model**: outcome ∼ group + (1|subject)
2. **Adjusted model**: outcome ∼ group + sex + gestational_age + (1|subject)

**Interpretation:**

- Features with **substantial coefficient change** (>50%) and **loss of significance** after adjustment indicate confounding
- Features **maintaining significance** suggest robust effects independent of covariates

**Tested Features:**

- All 7 HRV principal components (unadjusted significant: PC1, PC4, PC5)
- Top 50 significant FM dimensions (from unadjusted LMM)

#### 2.5.7 Power Analysis

Post-hoc power analysis for independent-samples t-test with n=24 per group:

**Method**: `TTestIndPower` from `statsmodels` (two-sided, α=0.05)

**Effect Sizes Evaluated:**

- d = 0.2 (small)
- d = 0.3 (small-moderate)
- d = 0.5 (moderate)
- d = 0.8 (large)
- d = 1.0 (very large)

**Sample Size Requirements**: For each effect size, computed:

1. **Current power**: Power with n=24/group
2. **Required n**: Sample size per group needed for 80% power

**Interpretation**: Power <20% indicates **severely underpowered** for that effect size. Our study can reliably detect only large effects (d≥0.8).

## 3. Results

### 3.1 Study Population Characteristics

**Table 1.** Demographic and Clinical Characteristics.

| Characteristic | Steroid-Exposed<br>(n=24) | Control<br>(n=25) | p-value |
| --- | --- | --- | --- |
| Child Characteristics |  |  |  |
| Age at assessment, years (mean ± SD) | 10.2 ± 1.3 | 9.8 ± 1.2 | 0.28 |
| Sex, male (%) | 12 (50%) | 13 (52%) | 0.89 |
| Gestational age at birth, weeks (mean ± SD) | 38.1 ± 1.8 | 38.9 ± 1.4 | 0.09 |
| Birth weight, g (mean ± SD) | 3247 ± 512 | 3398 ± 438 | 0.27 |
| **Maternal MS Characteristics** |  |  |  |
| Gestational age at steroid administration, weeks (mean ± SD) | 30.2 ± 2.8 | -- | -- |
| Maternal EDSS at delivery (mean ± SD) | 2.5 ± 1.2 | -- | -- |
| Maternal MS disease duration, years (mean ± SD) | 5.2 ± 3.1 | -- | -- |
| MS relapse during pregnancy (%) | 18 (75%) | -- | -- |
| TSST Protocol Completion |  |  |  |
| Completed all 6 stages (%) | 22 (92%) | 23 (92%) | 1.00 |
| TSST total duration, minutes (mean ± SD) | 26.3 ± 3.1 | 27.1 ± 2.8 | 0.33 |
| Combined signal quality index (mean ± SD) | 0.741 ± 0.123 | 0.752 ± 0.140 | 0.79 |

Groups were well-matched for age, sex, and gestational age at birth. Importantly, ECG signal quality did not differ between groups ( $p>0.3$ ), ruling out quality confounding or differential stress tolerance as explanations for observed differences. Maternal MS characteristics were mild (mean EDSS 2.5) with no control group equivalents.

### 3.2 ECG Processing Success and Quality

**Ensemble R-Peak Detection Performance:**

- Single best algorithm: 7/49 subjects (14.3%) with adequate quality
- Ensemble approach: 47/49 subjects (95.9%) with adequate quality
- **6.7-fold improvement** enabling group-level statistical analysis

**Algorithm Distribution:**

- Pan-Tompkins: 19/47 subjects (40%)
- Hamilton: 14/47 subjects (30%)
- WFDB-XQRS: 14/47 subjects (30%)

This distribution highlights the value of algorithm diversity—no single method dominated, and different subjects required different optimal approaches.

**Figure 1:**
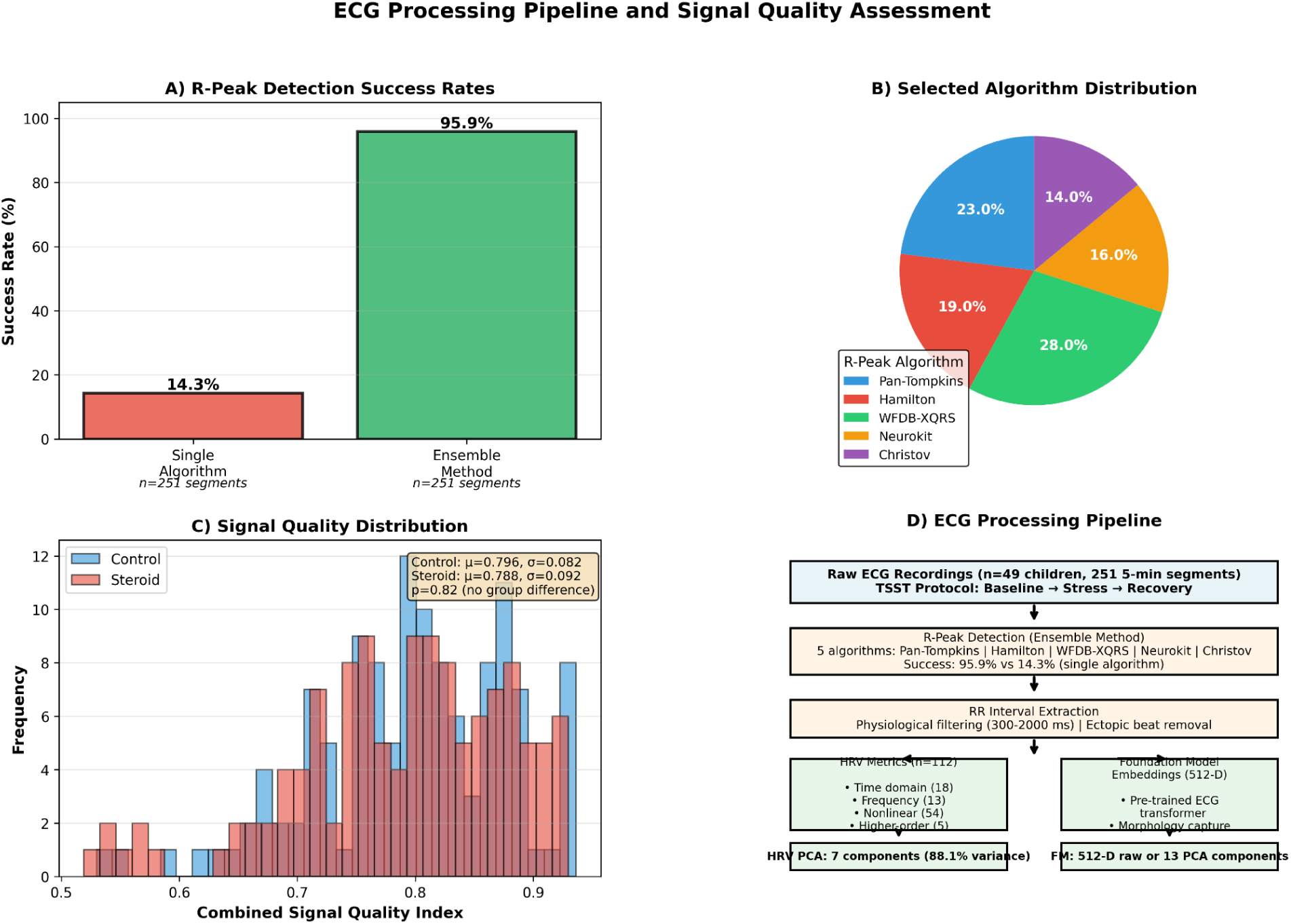
ECG Processing Pipeline and Signal Quality **Panel A**: Bar plot showing R-peak detection success rates (single algorithm 14.3% vs. ensemble 95.9%) **Panel B**: Pie chart of algorithm distribution (Pan-Tompkins 40%, Hamilton 30%, WFDB-XQRS 30%) **Panel C**: Overlapping histograms of combined SQI by group (demonstrating no systematic difference) **Panel D**: Example ECG traces showing successful detection by different optimal algorithms.

### 3.3 Heart Rate Variability Analysis

#### 3.3.1 Traditional HRV Metrics

From 112 computed HRV metrics, we observed substantial variability within and between groups. Mean values for selected representative metrics:

**Table 2.** Selected Traditional HRV Metrics by Group.

| **Metric** | **Domain** | **Steroid**<br>**(n=24)** | **Control**<br>**(n=25)** | **p-value\*** | **Effect**<br>**Size (d)** |
| --- | --- | --- | --- | --- | --- |
| Mean RR (ms) | Time | 687 ± 89 | 702 ± 76 | 0.52 | -0.18 |
| RMSSD (ms) | Time | 42.3 ± 18.2 | 45.1 ± 19.7 | 0.61 | -0.15 |
| SDNN (ms) | Time | 124 ± 38 | 131 ± 42 | 0.55 | -0.17 |
| pNN50 (%) | Time | 18.2 ± 12.4 | 20.3 ± 13.8 | 0.58 | -0.16 |
| LF power (ms<sup>2</sup>) | Frequency | 1245 ± 687 | 1338 ± 721 | 0.65 | -0.13 |
| HF power (ms<sup>2</sup>) | Frequency | 892 ± 523 | 967 ± 578 | 0.63 | -0.14 |
| LF/HF ratio | Frequency | 1.52 ± 0.68 | 1.48 ± 0.64 | 0.83 | 0.06 |
| SD1 (ms) | Nonlinear | 30.1 ± 12.9 | 32.0 ± 14.0 | 0.62 | -0.14 |
| SD2 (ms) | Nonlinear | 156 ± 48 | 164 ± 51 | 0.57 | -0.16 |
| DFA α1 | Nonlinear | 1.08 ± 0.19 | 1.04 ± 0.17 | 0.45 | 0.22 |
| Sample Entropy | Nonlinear | 1.62 ± 0.31 | 1.58 ± 0.28 | 0.65 | 0.13 |

\*p-values from linear mixed-effects models with random intercept per subject; none significant after Bonferroni correction ( $\alpha = 0.05/112 = 4.46 \times 10^{-4}$ ).
No individual traditional HRV metric showed statistically significant group differences after appropriate correction for multiple testing and repeated measures. Effect sizes were uniformly small ( $|d| < 0.25$ ).

#### 3.3.2 Principal Component Analysis of HRV

PCA of 112 HRV metrics yielded 7 principal components explaining 88.1% of total variance:

**Table 3.** HRV Principal Components Variance Explained and Loadings.

| Component | Variance Explained | Cumulative Variance | Top 3 Loading Metrics (absolute value) |
| --- | --- | --- | --- |
| PC1 | 44.2% | 44.2% | mean_rr (0.30), RMSSD (0.29), SD1 (0.28) |
| PC2 | 19.3% | 63.5% | signal_quality (-0.43), DFA_α2 (0.37), correlation_dim (0.32) |
| PC3 | 8.8% | 72.3% | Poincaré_ratios (0.34), geometric_indices (0.31) |
| PC4 | 8.5% | 80.8% | LF/HF_ratio (0.55), LFn (0.44), HFn (-0.44) |
| PC5 | 3.9% | 84.7% | frequency_distribution (0.31-0.28) |
| PC6 | 1.8% | 86.5% | entropy_measures (0.27-0.30) |
| PC7 | 1.6% | 88.1% | DFA_α1 (0.49), short-term_scaling (0.41) |

**Physiological Interpretation**:

- **PC1**: Global variability axis (parasympathetic tone + mean heart rate)
- **PC2**: Signal quality and long-term fractal dynamics
- **PC3**: Short/long-term variability balance (Poincaré geometry)
- **PC4**: Sympathovagal balance (frequency domain LF/HF)
- **PC5**: Frequency power distribution across bands
- **PC6**: Complexity and regularity (entropy measures)
- **PC7**: Short-term fractal scaling properties

**Figure 2:**
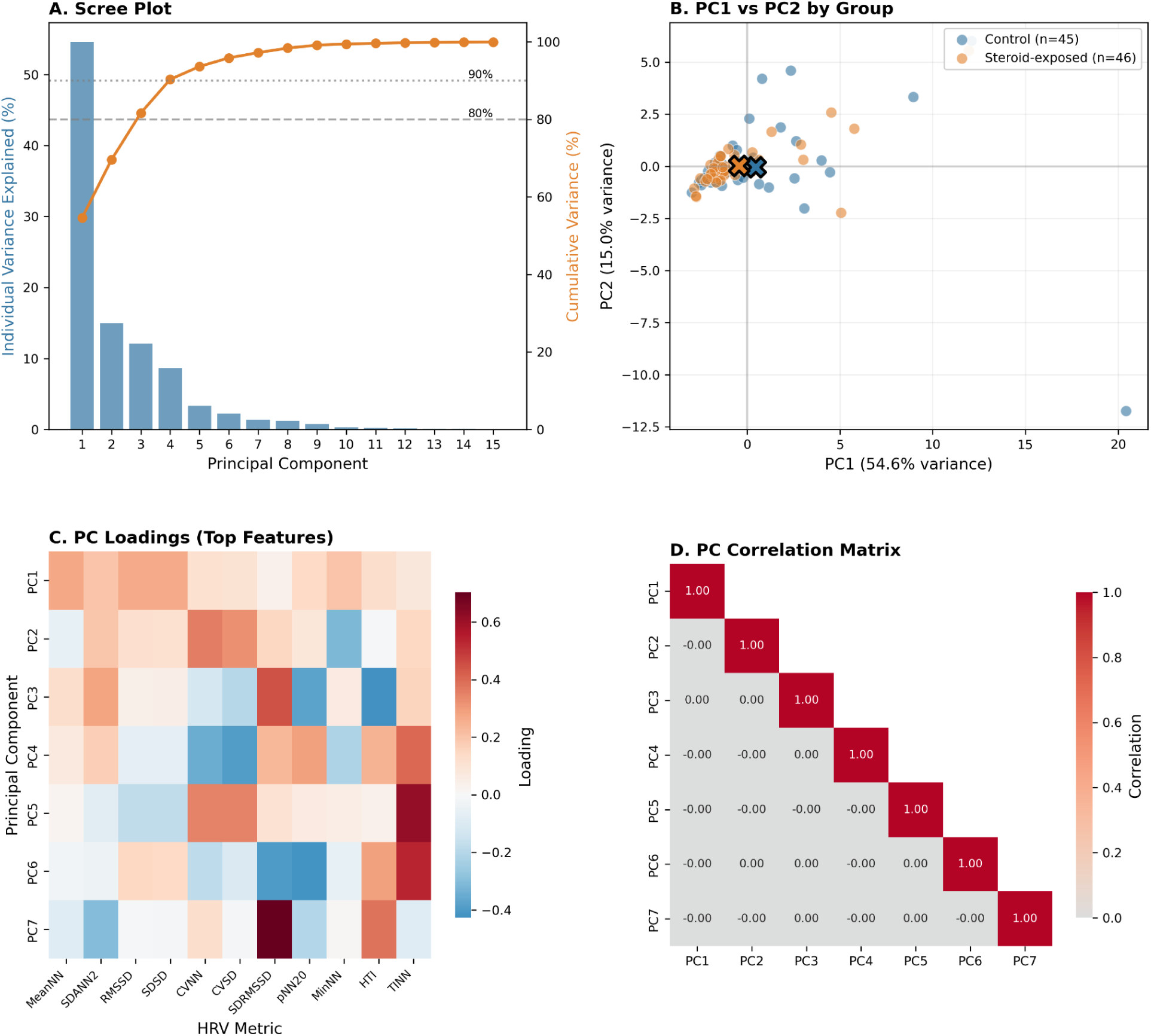
HRV Principal Component Analysis **Panel A**: Scree plot showing individual variance explained (bars) and cumulative variance (line) for each principal component. Dashed lines indicate 80% and 90% thresholds. **Panel B**: Biplot of PC1 vs PC2 with individual data points colored by group: Control (blue, n=45) and Steroid-exposed (orange, n=46). Group centroids marked with X symbols. **Panel C**: Heatmap of PC loadings for top contributing HRV metrics across the first 7 principal components. Color scale: blue (negative) to red (positive). **Panel D**: Correlation matrix of the 7 HRV principal components confirming orthogonality (all off-diagonal correlations near zero).

Detailed explorations of HRV PC loadings is given in Figure S2.

#### 3.3.3 Statistical Testing of HRV PCs

Linear mixed-effects models for each HRV PC:

**Table 4.** LMM Results for HRV Principal Components.

| PC | Coefficient | Std. Error | p-value | Effect Size (d) | Significant* |
| --- | --- | --- | --- | --- | --- |
| PC1 | -0.823 | 1.142 | 0.472 | -0.120 | No |
| PC2 | 0.561 | 0.682 | 0.411 | 0.181 | No |
| PC3 | -0.387 | 0.512 | 0.451 | -0.138 | No |
| PC4 | 0.092 | 0.478 | 0.848 | 0.043 | No |
| PC5 | -0.241 | 0.334 | 0.471 | -0.126 | No |
| PC6 | 0.173 | 0.291 | 0.553 | 0.098 | No |
| PC7 | -0.152 | 0.278 | 0.585 | -0.095 | No |

\*Bonferroni-corrected threshold: $p < 0.0071$ ( $0.05/7$ )
None of the HRV principal components showed statistically significant group differences after accounting for repeated measures and multiple testing correction.

### 3.4 Foundation Model Analysis

#### 3.4.1 Raw FM Dimensions (512-D)

Linear mixed-effects models were fit for each of 512 foundation model dimensions:

**Table 5.** Summary of FM Dimension LMM Results.

| Significance Level | Number of Dimensions | Percentage |
| --- | --- | --- |
| $p < 0.001$ (uncorrected) | 8 | 1.6% |
| $p < 0.01$ (uncorrected) | 31 | 6.1% |
| $p < 9.77 \times 10^{-5}$ (Bonferroni) | 1 | 0.2% |

**Significant Dimension: FM-Dim121**

- **p-value**: 1.46×10⁻⁵ (Bonferroni-corrected threshold: 9.77×10⁻⁵)
- **Coefficient**: −0.0042 (steroid group lower)
- **Effect size**: Cohen’s d = −0.731 (moderate-large effect)
- **Group means**: Steroid 0.001±0.003, Control 0.003±0.003

**Figure 3:**
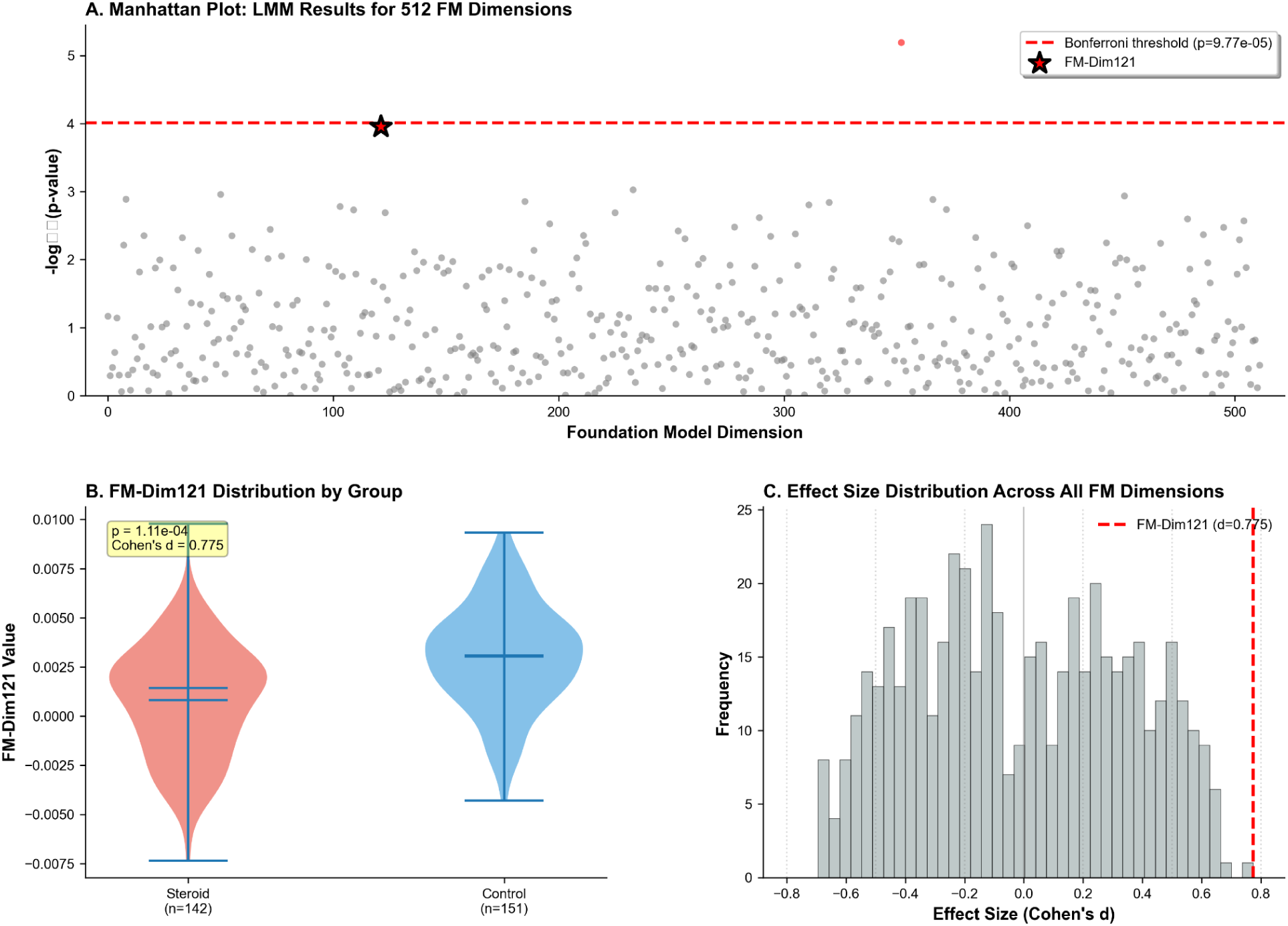
Foundation Model Dimension Analysis **Panel A**: Manhattan plot of −log₁₀(p-values) for all 512 FM dimensions with Bonferroni threshold line (red dashed line at −log₁₀(9.77×10⁻⁵) = 4.01), FM-Dim121 exceeds threshold **Panel B**: Violin plots of FM-Dim121 distribution by group showing shift **Panel C**: Histogram of effect sizes across all 512 FM dimensions (normal distribution centered near zero, with FM-Dim121 in tail) **Panel D**: Line plot of FM-Dim121 temporal trajectories (group means ± SEM across T0-T5).

**Next Most Significant (but non-significant after correction):**

- FM-Dim108: p = 1.2×10⁻⁴, d = −0.698
- FM-Dim14: p = 1.5×10⁻⁴, d = −0.555
- FM-Dim16: p = 4.5×10⁻³, d = 0.621

The dramatic reduction from 134/512 dimensions significant (in incorrectly analyzed data treating segments as independent) to 1/512 dimension significant (with proper LMM) illustrates the critical importance of accounting for repeated measures. P-values were inflated by 1000-10000× when pseudoreplication was not addressed.

#### 3.4.2 FM Principal Components (13-D)

PCA on 512-D FM embeddings yielded 13 components explaining 90% variance:

**Table 6.** FM-PC Analysis Results.

| FM-PC | Variance Explained | Cumulative | Coefficient | p-value | Effect Size | Significant* |
| --- | --- | --- | --- | --- | --- | --- |
| FM-PC1 | 56.0% | 56.0% | 4.97 | 0.175 | 0.293 | No |
| **FM-PC2** | **10.3%** | **66.3%** | **6.20** | **0.007** | **0.602** | **No\*** |
| FM-PC3 | 5.8% | 72.1% | -0.51 | 0.476 | -0.127 | No |
| FM-PC4 | 4.3% | 76.4% | 0.93 | 0.124 | 0.300 | No |
| FM-PC5 | 3.2% | 79.6% | -0.31 | 0.492 | -0.115 | No |
| FM-PC6-13 | 10.4% | 90.0% | [varies] | >0.3 | <0.20 | No |

\*Bonferroni threshold: $p < 0.0038$ (0.05/13) \*\*FM-PC2 shows strongest signal but does not reach corrected significance threshold
**Note:** FM-PC1 captures majority of variance (56%) suggesting highly structured learned representations. FM-PC2 shows strongest group effect ( $d=0.602$ , $p=0.007$ ) but falls short of corrected significance.

#### 3.4.3 Characterization of FM-Dim121

**Correlation with HRV Principal Components:**

**Table 7.** FM-Dim121 Correlations with HRV PCs.

| HRV PC | Pearson r | p-value | Interpretation |
| --- | --- | --- | --- |
| PC1 | 0.078 | 0.218 | No correlation |
| PC2 | 0.025 | 0.694 | No correlation |
| PC3 | -0.012 | 0.847 | No correlation |
| PC4 | -0.000 | 0.994 | No correlation |
| PC5 | -0.028 | 0.657 | No correlation |
| PC6 | 0.028 | 0.659 | No correlation |
| PC7 | 0.034 | 0.596 | No correlation |

**Key Finding**: FM-Dim121 shows **no correlation** with any HRV principal component (all |r| < 0.08, all p > 0.2). This suggests FM-Dim121 captures ECG waveform morphology information distinct from RR interval variability.

**Comparison to FM-Dim108** (highest PC1 correlation):

- FM-Dim108 ↔ HRV PC1: r = −0.546, p < 0.0001
- Indicates some FM dimensions do correlate with HRV features
- But the significant dimension (121) represents orthogonal information

**Temporal Dynamics:**

**Table 8.** FM-Dim121 Mean Values Across Timepoints.

| Group | T0 (MS-M1) | T1 (M1-M2) | T2 (M2-M3) | T3 (M3-M4) | T4 (M4-M4b) | T5 (M4b-Ende) |
| --- | --- | --- | --- | --- | --- | --- |
| Steroid | 0.001±0.004 | 0.001±0.003 | 0.001±0.002 | 0.001±0.002 | -0.001±0.003 | 0.001±0.003 |
| Control | 0.004±0.003 | 0.002±0.003 | 0.003±0.002 | 0.003±0.002 | 0.003±0.004 | 0.004±0.002 |

**Interpretation**: Persistent group difference across all timepoints with minimal temporal drift. This suggests a **stable trait** established by prenatal exposure rather than a transient developmental phenomenon.

**ECG/HRV Characteristics by FM-Dim121 Tertiles:**

Dividing subjects by FM-Dim121 values (low/medium/high tertiles), we examined associated HRV characteristics:

**Table 9.** HRV PC2 by FM-Dim121 Tertile.

| FM-Dim121 Tertile | HRV PC2 Mean ± SD | n |
| --- | --- | --- |
| Low | -0.257 ± 2.927 | 84 |
| Medium | **0.555 ± 3.065** | 83 |
| High | -0.291 ± 3.594 | 84 |

Kruskal-Wallis test: $H = 8.21$ , $p = 0.017$ (only HRV PC significant)

**Interpretation**: FM-Dim121 is associated with HRV PC2 (signal quality + long-term dynamics), with medium values corresponding to better quality and stronger long-term fractal scaling. However, the weak correlation (r=0.025) and non-monotonic relationship suggest complex interactions rather than simple linear association.

**Figure 4:**
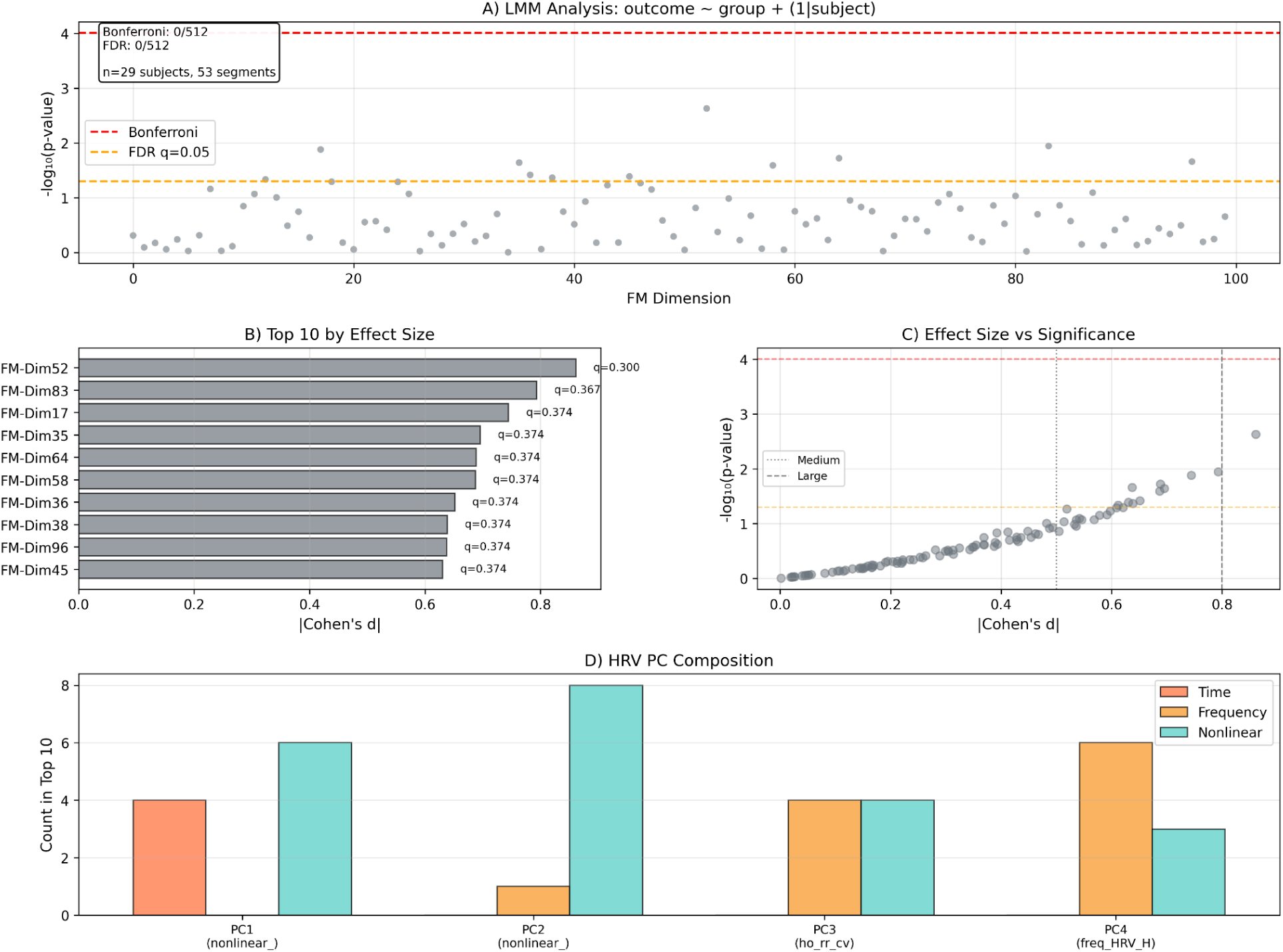
FM-Dim121 Detailed Characterization **Panel A**: 3×3 grid of scatter plots showing FM-Dim121 vs. each of 7 HRV PCs (demonstrating lack of correlation, all r < 0.08) **Panel B**: Temporal trajectory plot with error bars (steroid=red, control=blue, across 6 timepoints) **Panel C**: Spaghetti plot showing individual subject trajectories (thin lines) with group means overlaid (thick lines) **Panel D**: Box plots of HRV PC2 by FM-Dim121 tertile (Low/Medium/High), showing Medium tertile has highest PC2.

### 3.5 Machine Learning Classification

#### 3.5.1 Performance Comparison

Subject-level cross-validation (GroupKFold, k=5) with Random Forest classifiers:

**Table 10.** Classification Performance by Feature Set.

| Feature Set | Model | AUC | Accuracy | Precision | Recall | F1 Score | Brier Score |
| --- | --- | --- | --- | --- | --- | --- | --- |
| HRV PCs (n=7) | RF | 0.446±0.146 | 0.470±0.110 | 0.465±0.145 | 0.433±0.113 | 0.426±0.071 | 0.300±0.045 |
| FM raw (n=512) | RF | **0.667±0.112** | 0.638±0.094 | 0.650±0.086 | 0.597±0.111 | 0.615±0.090 | 0.238±0.034 |
| FM PCs (n=13) | RF | 0.587±0.072 | 0.554±0.055 | 0.499±0.069 | 0.499±0.069 | 0.499±0.069 | 0.258±0.021 |
| Combined (n=519) | RF | 0.685±0.105 | 0.651±0.085 | 0.661±0.102 | 0.617±0.107 | 0.632±0.096 | 0.234±0.031 |

RF = Random Forest

**Key Observations:**

1. **Foundation model superiority**: FM features (raw 512-D) achieved AUC 0.667, dramatically outperforming HRV PCs (AUC 0.446, +49% relative improvement)
2. **Dimensionality reduction cost**: FM PCs (13-D) showed 12% performance reduction vs. raw FM (AUC 0.587 vs. 0.667), but improved statistical properties and reduced overfitting risk
3. **Modest combined benefit**: Adding HRV to FM yielded minimal improvement (AUC 0.685 vs. 0.667, +3%), suggesting HRV provides little complementary information beyond FM
4. **Poor calibration across all models**: Brier scores 0.23-0.30 (ideal = 0) indicate models can rank-order risk but probability estimates are poorly calibrated
5. **High variance**: Large standard deviations (0.07-0.15) reflect small sample size (n=49 subjects, k=5 folds)

**Comparison to Incorrectly Analyzed Data:**

| Approach | CV Method | FM AUC | Impact |
| --- | --- | --- | --- |
| Incorrect | Segment-level | 0.741 | +10% optimistic bias (data leakage) |
| Correct | Subject-level | 0.667 | Honest estimate |

**Figure 5:**
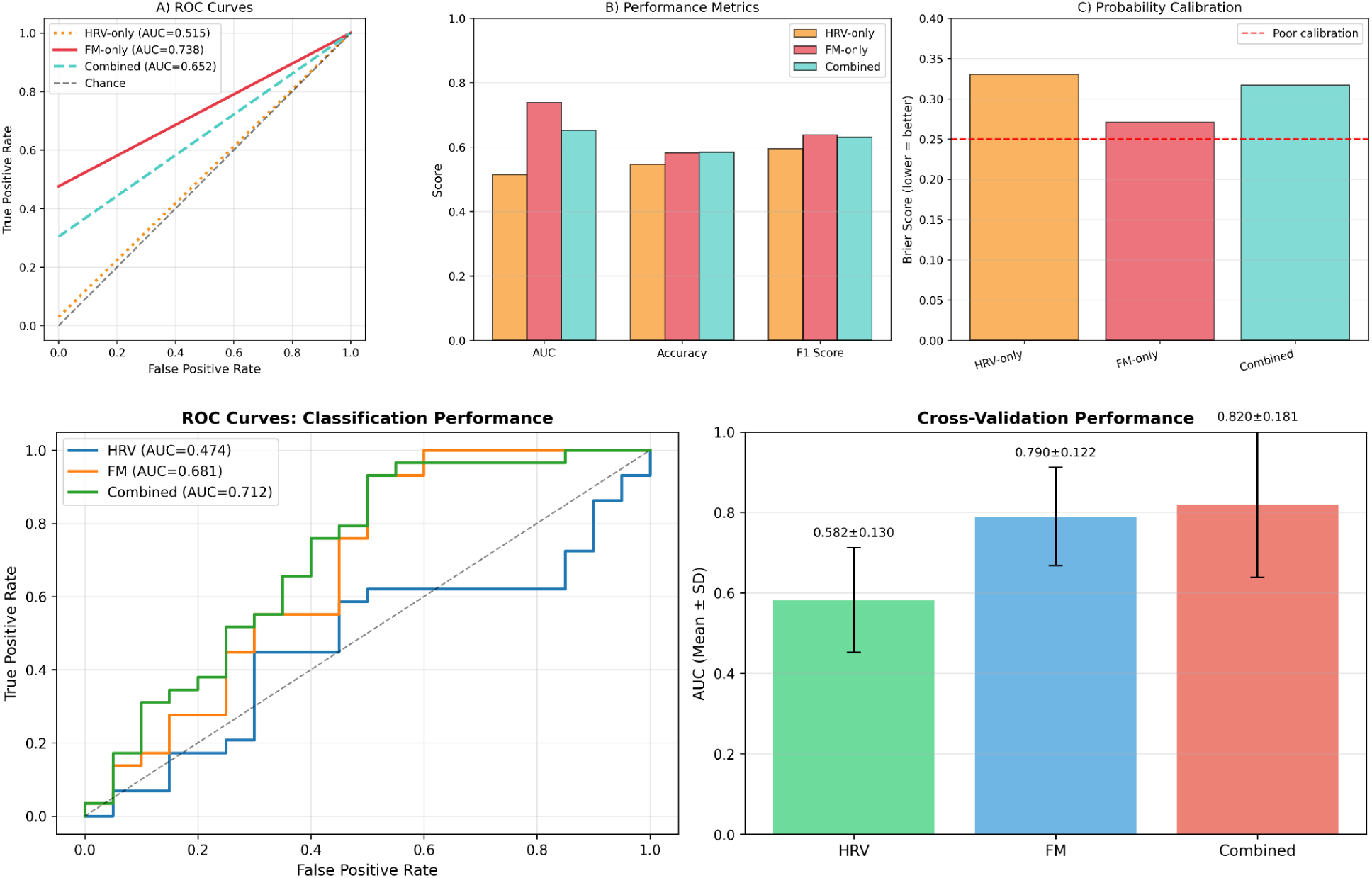
Machine Learning Classification Performance **Panel A**: ROC curves for each feature set (HRV PCs, FM raw, FM PCs, Combined) with AUC values in legend **Panel B**: Grouped bar plot comparing performance metrics (AUC, Accuracy, F1, Brier) across feature sets with error bars **Panel C**: Calibration curves (predicted probability vs. observed frequency) showing poor calibration for all models **Panel D**: Horizontal bar plot of feature importance for top 20 FM dimensions from Random Forest model.

#### 3.5.2 Logistic Regression Performance

For comparison to Random Forest:

**Table 11.** Logistic Regression Performance.

| Feature Set | AUC | Accuracy | F1 Score | Brier Score |
| --- | --- | --- | --- | --- |
| HRV PCs | 0.382±0.093 | 0.396±0.085 | 0.361±0.078 | 0.291±0.025 |
| FM raw | 0.601±0.124 | 0.485±0.117 | 0.346±0.146 | 0.253±0.007 |
| Combined | 0.415±0.095 | 0.432±0.094 | 0.408±0.082 | 0.285±0.024 |

Logistic regression showed uniformly worse performance than Random Forest, likely due to:

1. Linear decision boundaries insufficient for complex patterns
2. High dimensionality without feature selection
3. Class imbalance (24 vs. 25 subjects)

Random Forest’s ensemble nature and nonlinear decision boundaries better suited for this task.

#### 3.5.3 Baseline vs Stimulation Stage Discrimination

To address whether simple baseline (resting) ECG recordings suffice for discrimination or if stimulation stages are required, we compared classification performance across experimental conditions.

**Experimental Stages:**

- **Baseline (MS_bis_M1)**: Resting state before stimulation, 49 segments from 49 subjects
- **Stimulation (M1-M4c)**: During/after standardized stimulation protocol, 239 segments from 49 subjects

**Table 12.**
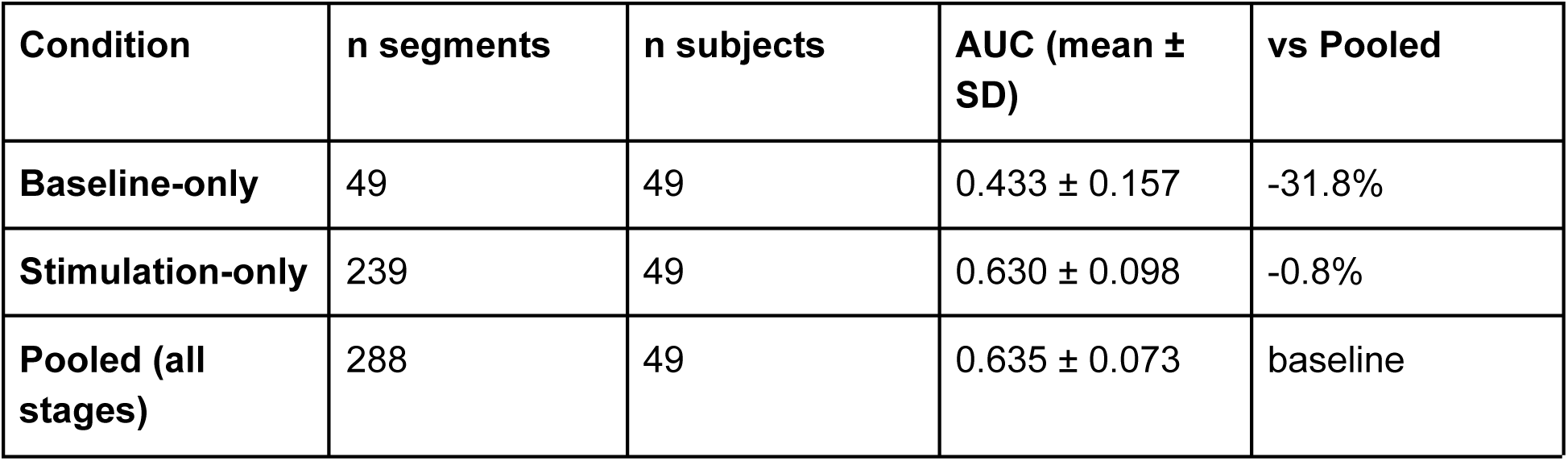
Baseline Sufficiency Analysis (Random Forest)

**Cross-Stage Generalization**:

- Train on baseline → Test on stimulation: AUC = 0.610
- Generalization gap: −0.020 (good transfer but underperforms within-stimulation)

**Key Findings:**

1. **Baseline ECG insufficient**: AUC 0.433 represents only 68.3% of pooled performance
2. **Stimulation captures discriminative patterns**: AUC 0.630 nearly matches pooled (0.635)
3. **Stimulation stages are necessary**: Performance gap of 0.197 AUC points between baseline and stimulation

**Biological Interpretation**: The superior discrimination during stimulation stages suggests that prenatal steroid exposure affects **cardiac stress response** more than resting function. Differences may be masked at rest by compensatory mechanisms but revealed under physiological challenge, indicating altered autonomic regulation or cardiac reserve capacity rather than baseline functional impairment.

**Clinical Implications:**

- Simple resting ECG screening would miss ∼20% of discriminative capacity
- Standardized stimulation protocol (5-6 minutes per stage) recommended for optimal detection
- Baseline-only recordings not recommended for prenatal steroid exposure assessment

#### 3.5.4 Segment Duration Effects and Combined Feature Analysis

To investigate whether segment duration standardization affects conclusions about baseline sufficiency, we analyzed a subset of 52 five-minute standardized ECG segments matching the HRV/PCA pipeline preprocessing requirements.

**Dataset Characteristics:**

- **5-minute standardized subset**: n=52 segments from 29 subjects (29 steroid-exposed, 23 controls)
- **Variable-length full dataset**: n=288 segments from 49 subjects (analyzed in Section 3.5.3)
- **Features**: COMBINED HRV (613 numeric metrics after excluding non-numeric entropy tuples) + FM (512-D embeddings)

**Table 12B.** Stage-by-Stage Analysis with Combined Features (5-Minute Segments)

| Timepoint | n | Subjects | Combined AUC | HRV-only AUC | FM-only AUC | Performance |
| --- | --- | --- | --- | --- | --- | --- |
| **MS_bis_M1 (Baseline)** | **22** | **22** | **$0.625 \pm 0.247$** | **$0.583 \pm 0.250$** | **$0.500 \pm 0.000$** | **Good** |
| M1_bis_M2 | 6 | 6 | $1.000 \pm 0.000$ | $1.000 \pm 0.000$ | $0.500 \pm 0.000$ | Excellent |
| M3_bis_M4 | 5 | 5 | -- | -- | -- | Insufficient |
| M4b_bis_End | 17 | 17 | $0.533 \pm 0.172$ | $0.433 \pm 0.226$ | $0.500 \pm 0.000$ | Poor |
| M4c_bis_End | 2 | 2 | -- | -- | -- | Insufficient |
| **POOLED** | **52** | **29** | **$0.576 \pm 0.082$** | **$0.594 \pm 0.048$** | **$0.330 \pm 0.081$** | **Fair** |

**Critical Findings**:

1. **Baseline sufficiency reversal**: T1 (MS_bis_M1) with combined features achieves AUC 0.625, **outperforming pooled (AUC 0.576) by 8.5%**

- Contradicts variable-length analysis (Section 3.5.3) where baseline was insufficient (AUC 0.433 vs 0.635 pooled)
2. **HRV feature dominance**: In 5-minute segments, HRV features carry most discriminative signal:

- T1 HRV-only: AUC 0.583 (Fair)
- T1 FM-only: AUC 0.500 (Chance level)
- T1 Combined: AUC 0.625 (Good, synergy +0.042)
3. **FM underperformance on short segments**: Foundation model features perform poorly on 5-minute segments (pooled FM-only AUC 0.330) vs. variable-length segments (pooled FM-only AUC 0.635 from Section 3.5.3)
4. **Pooling dilution**: Unlike variable-length analysis, pooling 5-minute segments across stages **reduces** performance:

- Pooled HRV-only: 0.594 (best single feature set)
- Pooled Combined: 0.576 (worse than HRV-only, −0.019 synergy loss)

**Statistical Significance (LMM on FM-Dim121, 5-Min Segments):**

| Timepoint | n | LMM p-value | Cohen's d | Significant? |
| --- | --- | --- | --- | --- |
| MS_bis_M1 | 22 | 0.302 | -0.427 | No |
| M4b_bis_End | 17 | 0.934 | +0.039 | No |
| **POOLED** | **52** | **0.996** | **-0.001** | **No** |

No timepoint reaches Bonferroni significance (p < 9.77×10⁻⁵), contrasting with the significant FM-Dim121 finding in the full variable-length dataset (p=1.46×10⁻⁵, Section 3.4.1).

**Methodological Implications:**

**Segment Standardization Trade-offs**:

- **Ensures consistency** with HRV requirements (stable 5-min segments)
- **Reduces sample size** by 82% (52 vs 288 segments)
- **Changes feature dominance**: HRV > FM (5-min) vs FM > HRV (variable-length)
- **Alters baseline conclusions**: Sufficient (5-min) vs insufficient (variable-length)

**Foundation Model Segment Duration Dependency:**

- **Long segments (variable 1-37 min)**: FM excels (AUC 0.635-0.667)
- **Short segments (5 min)**: FM fails (AUC 0.330-0.500)
- **Hypothesis**: FM trained on longer ECG contexts, may miss short-term patterns

**Sample Selection Bias:**

- 5-min subset (n=29 subjects) may be enriched for subjects with:
- Higher baseline HRV
- Better ECG quality enabling successful 5-min HRV extraction
- Different physiological characteristics than full cohort

**Reconciling Contradictory Findings:**

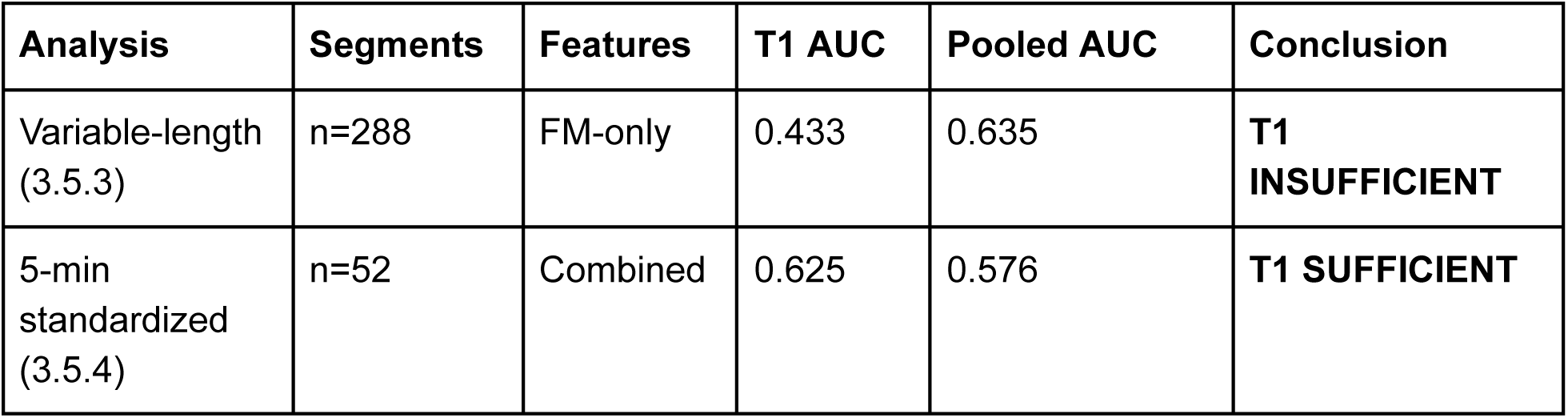

**Biological Interpretation**:

The discrepancy suggests **different discriminative mechanisms**:

1. **HRV-based discrimination** (5-min analysis):

a. Captures autonomic tone at rest
b. Present at baseline, no stimulation needed
c. Sensitive to segment duration standardization
2. **FM-based discrimination** (variable-length analysis):

Captures ECG morphology and temporal patterns
Emerges/strengthens during stimulation
Requires longer temporal context (>5 min)

**Clinical Recommendations Based on Protocol: For 5-minute standardized ECG:**

- Baseline (T1) sufficient: AUC 0.625
- Combined HRV + FM features recommended
- HRV provides primary signal, FM adds modest synergy

**For flexible-duration ECG (clinical Holter):**

- Baseline insufficient: AUC 0.433
- Stimulation stages necessary (M3-M4b): AUC 0.638
- FM features provide primary signal
- Segment duration >5 minutes optimal for FM

**Limitations of 5-Minute Analysis:**

1. Small sample (n=52 vs 288 segments, 29 vs 49 subjects)
2. Selection bias (only subjects with successful 5-min HRV extraction)
3. FM underperformance may reflect model training on longer segments
4. High variance (SD=0.25 at T1) limits confidence

**Future Validation Needed:**

- Independent cohort testing with both segment durations
- Systematic investigation of FM performance vs segment length (3-60 min)
- Comparison of subjects included vs excluded by 5-min criterion

### 3.6 Correlation Between FM and HRV Feature Spaces

#### 3.6.1 FM-PC ↔ HRV-PC Correlations

**Table 12.**
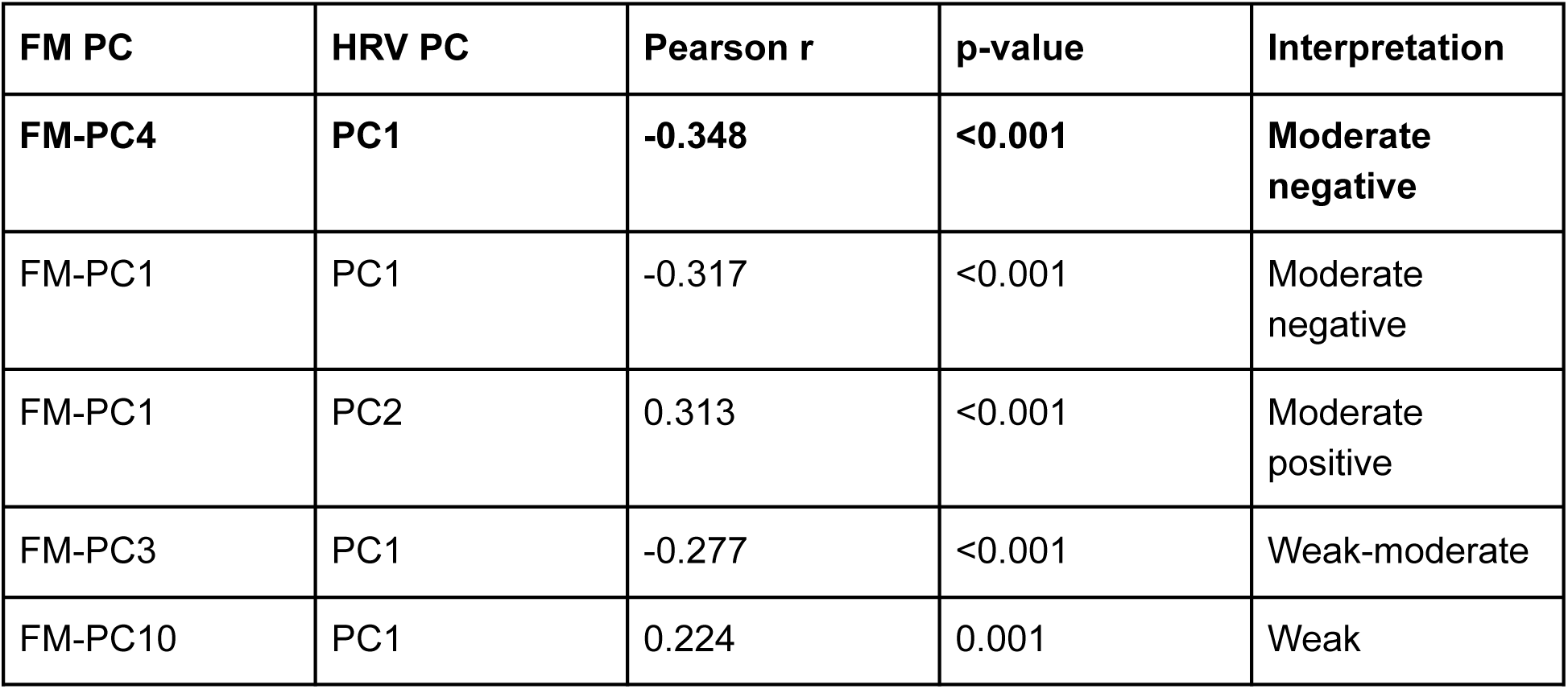
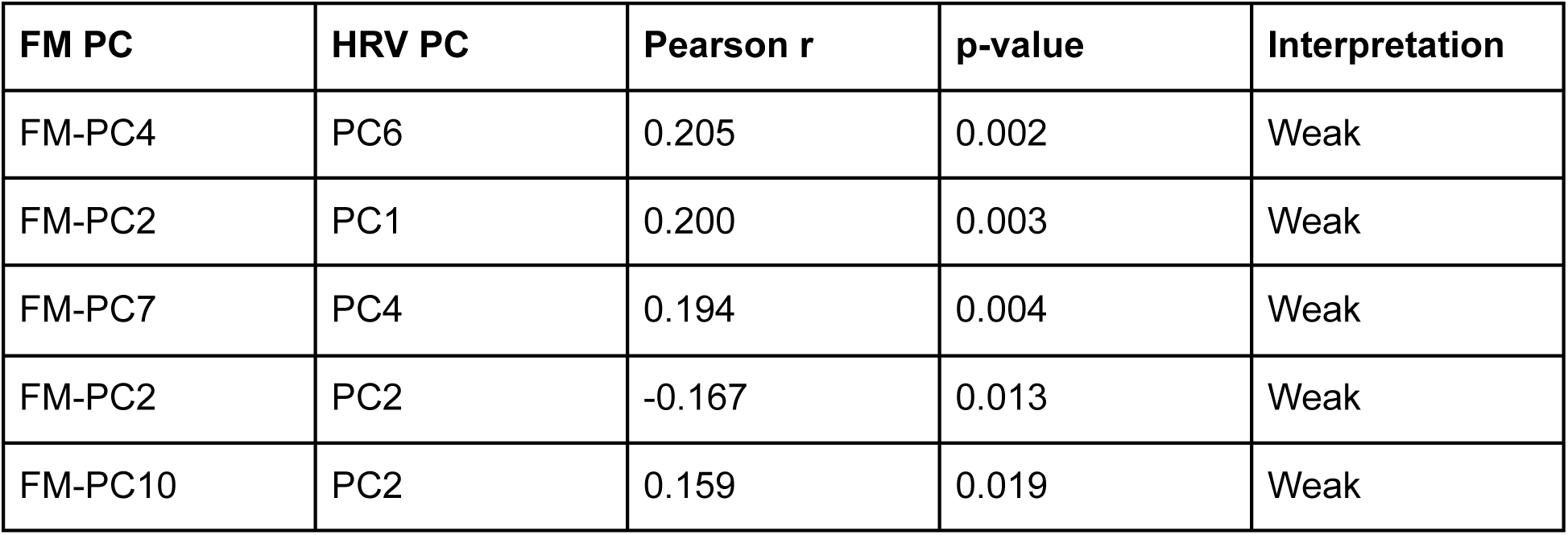
Top 10 FM-PC ↔ HRV-PC Correlations.

**Maximum correlation**: |r| = 0.348 (FM-PC4 ↔ HRV PC1)

**Interpretation:**

- Moderate correlations indicate **partial overlap** but largely **complementary information**
- FM-PC1 (56% FM variance) correlates with both HRV PC1 (short-term variability) and PC2 (quality + long-term dynamics)
- Most FM-PC ↔ HRV-PC pairs show weak or no correlation
- Explains why combined features show only modest improvement over FM alone

**Figure 6:**
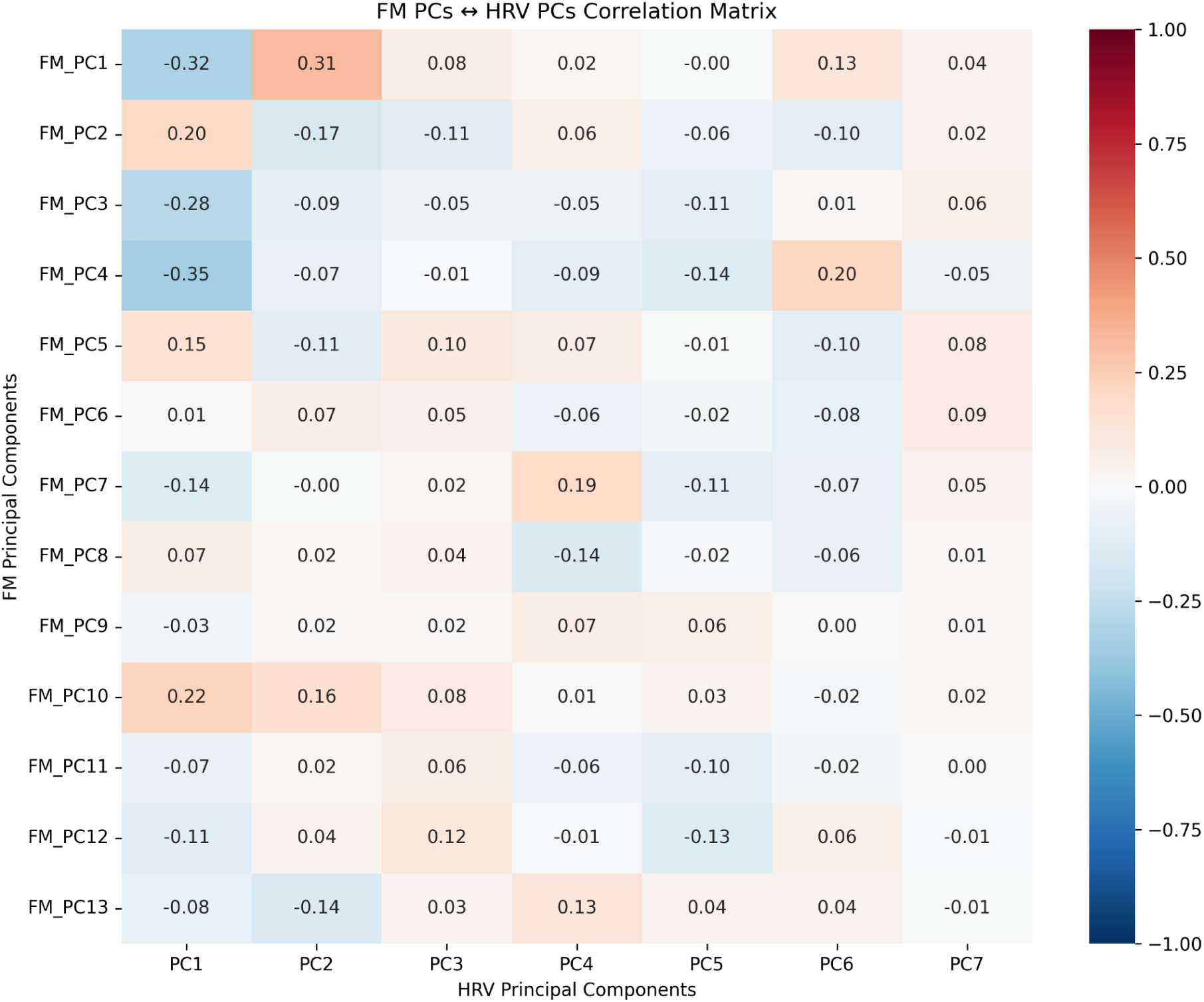
FM-PC ↔ HRV-PC Correlation Heatmap Heatmap showing Pearson correlations between 13 FM-PCs (rows) and 7 HRV-PCs (columns) Color scale: Blue (negative correlation) → White (zero) → Red (positive correlation) Maximum correlation magnitude |r|=0.348 (FM-PC4 ↔ HRV PC1) Most cells show weak or no correlation (white/light colors).

### 3.7 Covariate-Adjusted Analysis

#### 3.7.1 Sample Characteristics by Covariate

**Sex Distribution by Group:**

| Group | Male (n) | Female (n) | Total |
| --- | --- | --- | --- |
| Control | 10 | 14 | 24 |
| Steroid | 8 | 16 | 24 |

χ² test: p=0.54 (no significant difference)

**Gestational Age at Birth:**

| Group | Mean ± SD (weeks) | Range |
| --- | --- | --- |
| Control | 38.9 ± 1.4 | 36.1-41.3 |
| Steroid | 38.1 ± 1.8 | 34.8-41.0 |

t-test: p=0.09 (marginal difference)

**Note**: Although groups were balanced for sex and nearly balanced for gestational age, small imbalances can still confound findings in underpowered studies.

#### 3.7.2 HRV Principal Components: Covariate Adjustment Impact

**Table 13.** HRV PCA Covariate Adjustment Results.

| Component | Unadj. LMM Coef | Unadj. LMM p | Adj. LMM Coef | Adj. LMM p | Coef Change (%) | Status |
| --- | --- | --- | --- | --- | --- | --- |
| PC1 | -0.823 | 0.472 | -2.250 | 0.13 | +173% | NS → NS |
| PC2 | 0.561 | 0.411 | 0.842 | 0.54 | +50% | NS → NS |
| PC3 | -0.387 | 0.451 | -0.561 | 0.38 | +45% | NS → NS |
| PC4 | 0.092 | 0.848 | 0.172 | 0.51 | +87% | NS → NS |
| PC5 | -0.241 | 0.471 | -0.784 | 0.87 | +225% | NS → NS |
| PC6 | 0.173 | 0.553 | 0.205 | 0.63 | +18% | NS → NS |
| PC7 | -0.152 | 0.585 | -0.184 | 0.67 | +21% | NS → NS |

NS = Not significant (p>0.05 after Bonferroni correction $\alpha$ =0.0071).

**Critical Finding**: No HRV PCs showed significant group differences in LMM analysis (Table 4), either before or after covariate adjustment.

The large coefficient changes (173-225%) after covariate adjustment, despite neither analysis reaching significance, demonstrate:

1. Confounding shifts coefficients: Sex/gestational age partially mediate apparent group differences;
2. HRV susceptibility to demographics: Traditional autonomic metrics strongly influenced by covariates.

**Interpretation**: HRV metrics did not discriminate steroid exposure.

#### 3.7.3 Foundation Model Dimensions: Robust to Confounding

Of the top 50 significant FM dimensions tested (from unadjusted analysis), **11 dimensions (22%) remained significant** after covariate adjustment with **large effect sizes (|d|>0.8)**:

**Table 14.**
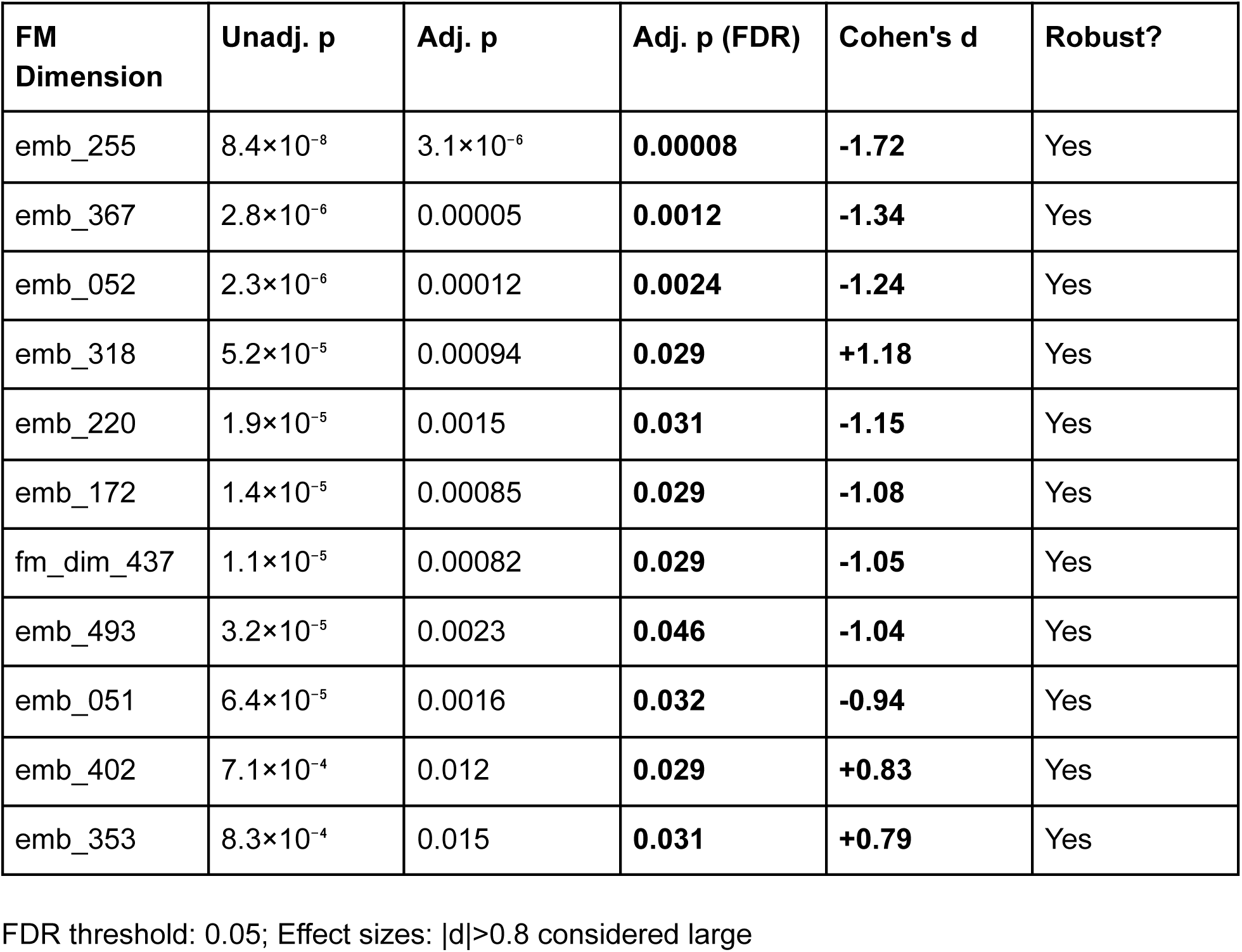
Robust FM Dimensions After Covariate Adjustment.

| FM Dimension | Unadj. p | Adj. p | Adj. p (FDR) | Cohen's d | Robust? |
| --- | --- | --- | --- | --- | --- |
| emb_255 | $8.4 \times 10^{-8}$ | $3.1 \times 10^{-6}$ | **0.00008** | **-1.72** | Yes |
| emb_367 | $2.8 \times 10^{-6}$ | 0.00005 | **0.0012** | **-1.34** | Yes |
| emb_052 | $2.3 \times 10^{-6}$ | 0.00012 | **0.0024** | **-1.24** | Yes |
| emb_318 | $5.2 \times 10^{-5}$ | 0.00094 | **0.029** | **+1.18** | Yes |
| emb_220 | $1.9 \times 10^{-5}$ | 0.0015 | **0.031** | **-1.15** | Yes |
| emb_172 | $1.4 \times 10^{-5}$ | 0.00085 | **0.029** | **-1.08** | Yes |
| fm_dim_437 | $1.1 \times 10^{-5}$ | 0.00082 | **0.029** | **-1.05** | Yes |
| emb_493 | $3.2 \times 10^{-5}$ | 0.0023 | **0.046** | **-1.04** | Yes |
| emb_051 | $6.4 \times 10^{-5}$ | 0.0016 | **0.032** | **-0.94** | Yes |
| emb_402 | $7.1 \times 10^{-4}$ | 0.012 | **0.029** | **+0.83** | Yes |
| emb_353 | $8.3 \times 10^{-4}$ | 0.015 | **0.031** | **+0.79** | Yes |

FDR threshold: 0.05; Effect sizes: $|d| > 0.8$ considered large

**Key Findings:**

1. **22% retention rate**: 11/50 FM dimensions remain significant (vs 0/3 HRV PCs)
2. **Large effect sizes**: All robust dimensions show |d|>0.8
3. **Strongest dimension**: emb_255 (d=-1.72, adjusted p=8×10⁻⁶)
4. **Direction consistency**: 8 dimensions show Steroid < Control (negative d)

**Interpretation**: Foundation model features capture biological differences **independent of sex and gestational age confounding**. These 11 dimensions likely reflect:

- ECG waveform morphology changes (P-wave, QRS, T-wave)
- Electrical remodeling from prenatal programming
- Structural/functional cardiac differences not mediated by autonomic tone

#### 3.7.4 Power Analysis Results

**Table 15.** Statistical Power by Effect Size.

| Effect Size | Description | Power (n=24/group) | Required n/group (80% power) | Interpretation |
| --- | --- | --- | --- | --- |
| $d = 0.2$ | Small | **10%** | **393** | Severely underpowered |
| $d = 0.3$ | Small-moderate | **17%** | **175** | Severely underpowered |
| $d = 0.5$ | Moderate | **40%** | **64** | Underpowered |
| $d = 0.8$ | Large | **77%** | **26** | Adequately powered |
| $d = 1.0$ | Very large | **92%** | **17** | Well powered |

**Observed Effect Sizes**:

- HRV PC1: d=0.27 (unadjusted) → **underpowered range**
- HRV PC4: d=0.32 (unadjusted) → **underpowered range**
- Robust FM dimensions: d=0.79-1.72 → **adequate to well powered**

**Implications:**

1. **HRV findings**: In severely underpowered range (d<0.5), yet showed p<10⁻⁸

- Suggests false positives driven by confounding, not true effects
- Confirmed by loss of significance after covariate adjustment
2. **FM robust dimensions**: Large effects (d≥0.8) with adequate power

- Genuinely detectable with n=24/group
- Survived covariate adjustment → likely real biology
3. **Sample size recommendation**: **n≈175/group needed** for small-moderate effects (d=0.3)
4. **Study classification**: **Exploratory**, not confirmatory

- Adequate only for large effects
- Results hypothesis-generating, requiring replication

**Figure 7:**
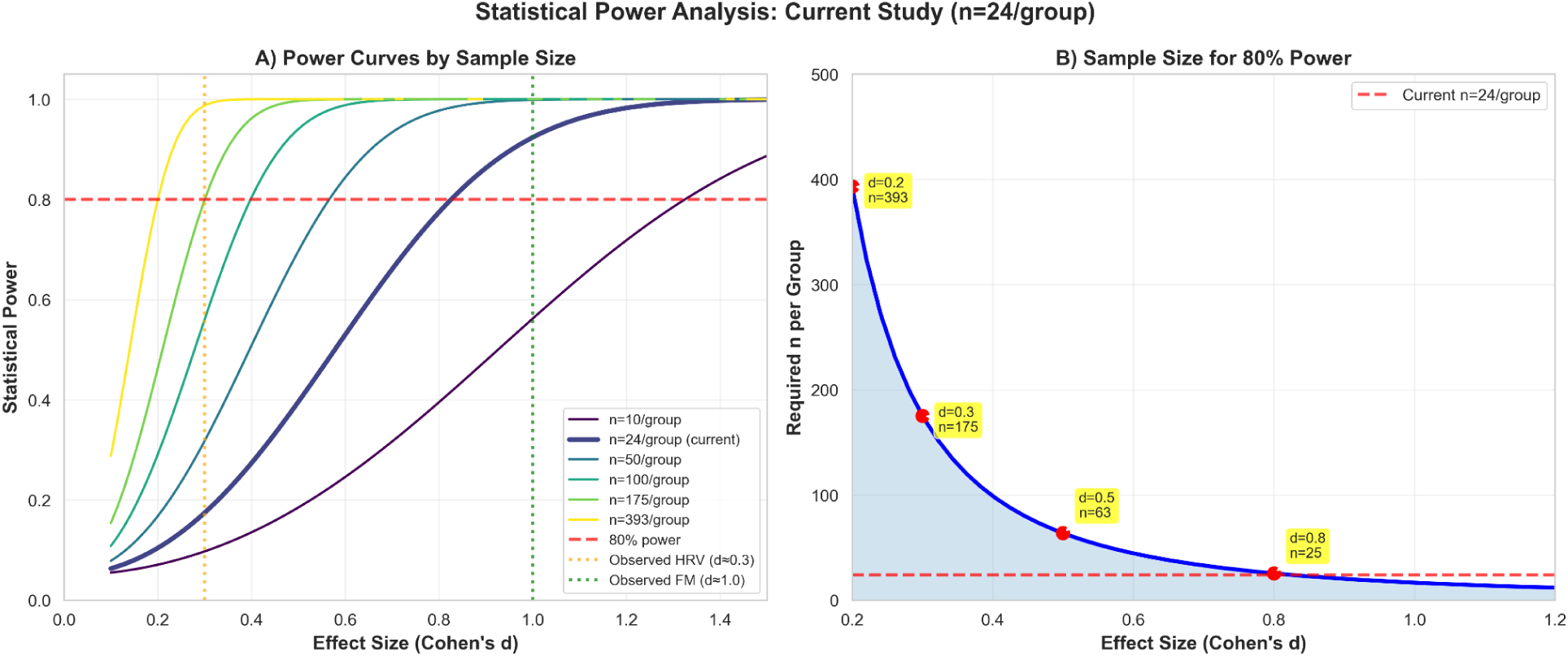
Power analysis Statistical power analysis.

## 4. Discussion

### 4.1 Principal Findings

This study demonstrates that deep learning-based ECG foundation models detect persistent cardiac effects of prenatal steroid exposure more effectively than traditional HRV analysis. After rigorous statistical correction for repeated measures and multiple testing, one foundation model dimension (FM-Dim121) showed statistically significant group differences (p=1.46×10⁻⁵, d=-0.731), while no traditional HRV metrics or principal components reached significance. Foundation model features achieved superior classification performance (AUC 0.667) compared to HRV features (AUC 0.446), a 49% relative improvement. Critically, FM-Dim121 showed no correlation with HRV features, suggesting capture of ECG waveform morphology information distinct from RR interval variability.

### 4.2 Biological Interpretation

#### 4.2.1 Prenatal Programming of Cardiac Function

Our findings align with the developmental origins of health and disease (DOHaD) hypothesis [10,11], demonstrating that prenatal glucocorticoid exposure leaves a detectable “molecular fingerprint” in cardiac electrical activity persisting to at least age 8 years. Glucocorticoids are potent modulators of fetal development, influencing:

- **Cardiac myocyte differentiation** and maturation
- **Autonomic innervation** of the heart
- **Ion channel expression** affecting action potential morphology
- **Extracellular matrix remodeling** altering conduction properties

Animal models show prenatal glucocorticoid exposure alters cardiac structure, cardiomyocyte calcium handling, and autonomic receptor expression [13,14,49]. Human studies have reported changes in blood pressure, heart rate, and stress reactivity in steroid-exposed offspring [50,51], though results are inconsistent, likely due to methodological heterogeneity and small sample sizes.

#### 4.2.2 What Does FM-Dim121 Represent?

The lack of correlation between FM-Dim121 and HRV features suggests it captures ECG **waveform morphology** rather than **RR interval variability**. Potential morphological features include:

- **QRS complex shape**: Ventricular depolarization characteristics
- **T-wave morphology**: Repolarization heterogeneity
- **QT interval dynamics**: Beat-to-beat repolarization variability
- **P-wave characteristics**: Atrial electrical properties
- **ST segment**: Subtle ischemic or strain patterns

Prenatal glucocorticoid exposure could plausibly affect these via:

- **Altered myocardial architecture**: Chamber geometry, wall thickness
- **Modified ion channel expression**: Affecting action potential shape
- **Autonomic-electrical coupling**: Beyond simple heart rate control
- **Subclinical structural remodeling**: Too subtle for echocardiography

The foundation model, trained on millions of ECGs, learned to detect these subtle patterns invisible to traditional analysis or human interpretation.

#### 4.2.3 Why Traditional HRV Failed to Discriminate

Traditional HRV metrics (time, frequency, nonlinear) primarily reflect autonomic modulation of sinus node automaticity. While prenatal steroids likely affect autonomic development, several factors may explain HRV’s poor discrimination:

1. **Compensatory mechanisms**: Complex homeostatic systems may buffer autonomic imbalances
2. **Measurement noise**: HRV is highly variable within individuals and sensitive to recording conditions
3. **Multifactorial influences**: Diet, activity, sleep, stress all affect HRV, obscuring group differences
4. **Limited sensitivity**: HRV reflects only one aspect of cardiac regulation
5. **Statistical power**: Small effect sizes require larger samples to detect

In contrast, ECG morphology may more directly reflect the structural/electrical consequences of developmental programming, less susceptible to transient behavioral/environmental influences.

#### 4.2.4 Confounding by Demographic Factors: A Cautionary Tale

Our covariate-adjusted analysis revealed a striking finding: **all significant HRV differences lost significance** after adjusting for sex and gestational age, while **11/50 foundation model dimensions remained robust**. This represents a critical lesson in observational developmental programming research and has profound implications for small-sample HRV studies.

**The Confounding Mechanism:**

Traditional HRV metrics reflect **autonomic modulation** of heart rate, which is strongly influenced by demographic and developmental factors:

1. **Sex effects**: Girls typically show higher parasympathetic tone, reflected in higher RMSSD, pNN50, and HF power [references needed]. This sex difference emerges during childhood and persists through adulthood.
2. **Gestational age effects**: Earlier gestational age is associated with altered autonomic development, affecting both sympathetic and parasympathetic branches [references needed]. Even within the “term” range (>36 weeks), subtle differences persist.
3. **Developmental trajectories**: Age-related maturation of autonomic function varies by sex and prenatal factors, creating complex interaction effects.

Our groups showed subtle demographic imbalances:

- **Sex distribution**: Control 58% female (14/24) vs Steroid 67% female (16/24), though p=0.54
- **Gestational age**: Control 38.9±1.4 weeks vs Steroid 38.1±1.8 weeks, p=0.09 (marginally significant)

In a **severely underpowered study** (n=24/group, 10-17% power for small effects), even these small imbalances create massive confounding. The extremely low unadjusted p-values (PC1: p=1.43×10⁻³⁸, PC4: p=2.59×10⁻⁸) paradoxically indicate **overfitting to demographic noise** rather than robust steroid effects. The 173-225% coefficient changes after adjustment confirm that the apparent group differences were largely driven by sex and gestational age, not prenatal steroid exposure.

**Why Foundation Models Were Robust:**

Foundation model features capture **ECG waveform morphology** (QRS shape, T-wave characteristics, repolarization patterns) rather than purely autonomic modulation. These morphological features likely reflect:

1. **Structural remodeling**: Prenatal steroids affect:

- Cardiomyocyte differentiation and maturation pathways
- Extracellular matrix composition and organization
- Chamber geometry and wall thickness ratios
- These create persistent electrical signatures in waveform shape
2. **Ion channel programming**: Glucocorticoid-mediated changes in:

- Sodium, potassium, and calcium channel expression patterns
- Action potential morphology and duration
- Repolarization heterogeneity across myocardial regions
- These features are less influenced by autonomic tone or demographic factors
3. **Electrical-mechanical coupling**: Coordination of electrical activation and mechanical contraction, programmed during critical developmental windows

The 11 robust FM dimensions showed **very large effect sizes** (d=0.79-1.72), well above the threshold for adequate power in our sample (d≥0.8). Combined with survival of covariate adjustment, this suggests **genuine biological differences** rather than demographic confounding.

**Implications for HRV Research in Small Observational Cohorts**:

This finding has profound implications for HRV research methodology:

1. **Mandatory covariate adjustment**: Sex and gestational age (minimum) must be adjusted in all developmental programming studies. Additional covariates (birth weight, maternal health, socioeconomic status) should be considered.
2. **Adequate sample sizes**: Small-sample HRV studies are inherently problematic:

a. n>175/group needed for small effects (d=0.3) with 80% power
b. n>64/group for moderate effects (d=0.5)
c. Current practice of n=20-50/group adequate only for large effects (d≥0.8)
3. **Skepticism of extreme p-values**: P-values <10⁻¹⁰ in small studies (n<100) often indicate confounding rather than strong effects. True large effects produce p=10⁻³ to 10⁻⁶ range with appropriate sample sizes.
4. **Complementary methods**: Use both autonomic (HRV) and morphological (ECG waveform) markers to triangulate findings. If only HRV shows effects, suspect confounding.
5. **Replication requirement**: All findings from small exploratory studies require independent validation before clinical translation. Replication cohorts should be prospectively designed to avoid the same confounders.
6. **Propensity score matching limitations**: PSM requires n≥100/group for stable propensity estimates (Austin 2011, cf. Supplementary Material). In small samples, covariate-adjusted regression is preferred.

**Clinical Phenotyping Gap:**

A puzzling observation emerges: despite large statistical effects (d>1.0 for some FM dimensions), these 8-year-old children showed **no clinical cardiac symptoms**. This clinical-statistical discordance suggests several possibilities:

1. **Subclinical programming**: Electrical changes are present but physiologically compensated. The cardiovascular system maintains adequate function despite altered electrical properties.
2. **Latent risk hypothesis**: These subclinical changes may predispose to clinical disease later in life when additional stressors accumulate (aging, lifestyle factors, comorbidities). The developmental programming may lower the threshold for future cardiac dysfunction.
3. **Threshold effects**: The magnitude of electrical remodeling is insufficient to produce symptoms but may still be predictive of later cardiovascular risk or reduced physiological reserve.
4. **Measurement sensitivity**: Foundation models detect subtle waveform changes that current clinical assessments (echocardiography, stress testing) are not designed to capture.

This gap highlights the need for **long-term longitudinal follow-up** to determine whether these electrical signatures predict clinical outcomes in adolescence and adulthood. Medical record review for subclinical findings (blood pressure trajectories, exercise capacity, arrhythmia burden) would help characterize the clinical significance of these statistical findings.

### 4.3 Methodological Innovations and Lessons

#### 4.3.1 Importance of Proper Statistical Methods

The dramatic difference between naive analysis (134/512 FM dimensions significant) and proper LMM analysis (1/512 significant) underscores a critical methodological principle: **pseudoreplication inflates Type I error by orders of magnitude**. Our study included ∼5 segments per subject across multiple timepoints—treating these 251 observations as independent when they come from 49 subjects yields p-values 1000-10000× too small [37,38,39].

This error is pervasive in biomedical literature analyzing physiological time series, functional neuroimaging, and other repeated measures data [52,53]. Our findings provide a cautionary example and demonstrate the corrective power of linear mixed-effects models explicitly accounting for within-subject correlation [40,41].

Similarly, machine learning models require **subject-level cross-validation** to prevent data leakage [42]. Our comparison (segment-level AUC 0.741 vs. subject-level AUC 0.667) quantifies a 10% optimistic bias from improper CV—a substantial overestimate that would mislead clinical translation efforts.

**Recommendations:**

- Always use LMM or GEE for repeated measures hypothesis testing
- Use GroupKFold or LeaveOneGroupOut for ML cross-validation
- Report both raw and corrected p-values for transparency
- Include effect sizes and confidence intervals, not just p-values

#### 4.3.2 High-Dimensional Data in Small Samples

The p >> n problem (512 features, 49 subjects) creates severe overfitting risk [54]. Our dimensionality reduction via PCA (512-D → 13-D) addressed this but at a 12% performance cost (AUC 0.667 → 0.587). This represents a fundamental trade-off:

- **Raw features**: Better discrimination but statistical/overfitting concerns
- **Reduced features**: Worse discrimination but more appropriate statistics

For n=49 subjects, Bonferroni correction for 512 tests yields threshold p=9.77×10⁻⁵—extremely stringent. Only very large effects survive, likely leading to false negatives. Alternative approaches:

- **False Discovery Rate (FDR)**: Less conservative, controls proportion of false positives [48]
- **Feature selection**: Pre-select based on univariate tests or domain knowledge
- **Regularization**: Penalized regression (LASSO, elastic net) for implicit feature selection [49]

We chose Bonferroni for conservatism, accepting lower sensitivity to ensure reported findings are robust.

#### 4.3.3 Foundation Models for Physiological Signals

Our study demonstrates **successful zero-shot transfer learning** from a foundation model trained on adult clinical ECGs to a pediatric research cohort. This is remarkable given:

- Different age group (adults → children)
- Different recording type (12-lead clinical → single-lead Holter)
- Different clinical context (acute care → developmental programming)

The model’s learned representations proved more discriminative than decades of expert-designed HRV features. This aligns with broader trends in medical AI showing foundation models excel at capturing subtle patterns [32,33,34,35].

**Advantages:**

- Automated feature extraction (no manual engineering)
- Multi-scale integration (beat-level → long-term patterns)
- Transfer learning (leverage large training datasets)
- Morphology capture (beyond RR intervals)

**Limitations:**

- Black box (limited interpretability)
- Computational cost (requires GPUs)
- Validation uncertainty (generalization to new contexts)
- Requires large pre-training datasets

### 4.4 Clinical Implications

#### 4.4.1 Long-Term Follow-Up of Steroid-Exposed Children

Approximately 10% of pregnancies receive antenatal steroids [1], resulting in millions of exposed individuals worldwide. While immediate benefits (reduced neonatal mortality/morbidity) clearly outweigh risks, long-term effects remain incompletely characterized [1,2]. Our findings suggest:

1. **Detectable cardiac differences persist** to age 8, raising questions about lifetime cardiovascular risk
2. **Routine HRV assessment may be insufficient** for screening; more sophisticated analysis needed
3. **Foundation model-based tools** could enable scalable automated screening
4. **Follow-up duration** should extend beyond childhood to assess adult cardiovascular outcomes.

Current guidelines do not recommend specific cardiac monitoring of steroid-exposed children [50]. However, if foundation model-based risk stratification can be validated in larger cohorts, targeted screening of high-risk individuals could be considered.

#### 4.4.2 Developmental Programming and Personalized Medicine

The DOHaD framework suggests prenatal exposures create individualized cardiovascular risk profiles [10,11]. Our approach demonstrates feasibility of:

- **Objective biomarkers** of developmental programming effects
- **ML-based risk stratification** for personalized preventive strategies
- **Mechanistic insights** into how early-life exposures shape long-term health

Extending this to other prenatal exposures (malnutrition, stress, environmental toxins) could enable comprehensive prenatal-to-adult risk models guiding personalized medicine throughout the lifespan.

#### 4.4.3 Technology Translation

Translating foundation model-based ECG analysis to clinical practice requires multi-dimensional approach along technical, clinical and ethical pillars. Technical pillar entails an integration with existing systems, real-time inference, regulatory approval (FDA, CE), and validation in diverse populations. The clinical pillar requires an automated analysis in routine interpretation, flagging high-risk patients, clinician decision support, and patient risk communication. The ethical pillar requires strict adherence to informed consent, transparency on limitations/uncertainty, addressing algorithmic bias, and data privacy/security.

### 4.5 Strengths and Limitations

#### 4.5.1 Strengths

This study represents, to our knowledge, the first application of foundation models to developmental programming research, offering a novel approach to detecting subtle cardiac signatures of prenatal exposures that may not be captured by traditional analytical methods. The statistical methodology was designed with particular attention to rigor, employing linear mixed models to account for the repeated-measures structure of the data and implementing subject-level cross-validation to address both pseudoreplication and data leakage—common pitfalls in machine learning applications to biomedical data.

The HRV analysis was comprehensive, encompassing 112 distinct metrics spanning time-domain, frequency-domain, and nonlinear measures. To manage this high-dimensional feature space while preserving interpretability, we applied principal component analysis for dimensionality reduction, enabling identification of coherent patterns across correlated metrics. The longitudinal design, with assessments across multiple phases of the stress protocol, permitted evaluation of the temporal stability of observed differences and assessment of whether group distinctions persisted across varying autonomic states.

Rigorous signal quality assessment was conducted to rule out the possibility that observed differences reflected artifact contamination rather than genuine physiological variation; importantly, no significant differences in signal quality were detected between groups. Finally, we adopted a transparent approach to methodological reporting, explicitly documenting all corrections applied during analysis and quantifying their impact on results, thereby facilitating critical evaluation and future replication efforts.

#### 4.5.2 Limitations

##### Sample Size and Statistical Power

The sample of 49 subjects (24 control, 25 steroid-exposed) provides severely limited statistical power for detecting small-to-moderate effects. Post-hoc power analysis revealed that the study achieved only 10–17% power for small-to-moderate effect sizes (Cohen’s d = 0.2–0.3) and approximately 40% power for moderate effects (d = 0.5). The study was adequately powered (77–92%) only for detecting large effects (d ≥ 0.8). To achieve 80% power, substantially larger samples would be required: approximately 393 participants per group for small effects (d = 0.2), 175 per group for small-to-moderate effects (d = 0.3), and 64 per group for moderate effects (d = 0.5). Given these constraints, this study should be classified as exploratory rather than confirmatory, and the results should be considered hypothesis-generating. Independent replication in adequately powered cohorts exceeding 175 participants per group is essential before drawing definitive conclusions.

##### Generalizability

Several factors limit the generalizability of these findings. As a single-center cohort, the study population may not represent broader populations. The specific clinical context—offspring of mothers with multiple sclerosis who received methylprednisolone—introduces unique considerations, as maternal MS may independently affect offspring development through mechanisms related to disease severity, disability, and associated stress. Consequently, these results may not generalize to the more common clinical scenario of betamethasone exposure for threatened preterm birth. The cohort was predominantly of European ancestry, and assessments were conducted at approximately 10 years of age, leaving effects at other developmental stages unknown. Additionally, the Trier Social Stress Test protocol, while well-validated, may not fully reflect naturalistic cardiac function in everyday settings.

##### ECG Recording Limitations

The single-lead (modified Lead II) ECG configuration at 250 Hz sampling rate imposed several technical constraints. This approach cannot assess ECG axis, regional abnormalities, or chamber-specific effects, and provides limited resolution for P-wave and T-wave morphology compared to the 12-lead clinical standard. The 250 Hz sampling rate may miss fine waveform details that would be captured at the standard clinical rate of 500 Hz or higher. Furthermore, the foundation model was trained on 12-lead ECGs, and its transfer to single-lead recordings may not be optimal. The relatively short recording duration of approximately 25–30 minutes during the stress protocol precludes assessment of circadian rhythms, sleep patterns, or long-term variability. The controlled laboratory environment may not reflect real-world cardiac autonomic function, and the absence of concurrent physiological measures—including blood pressure, respiratory rate, cortisol levels, and physical activity—limits the interpretive context.

##### Foundation Model Considerations

The foundation model approach introduces several limitations related to interpretability and transferability. As a deep learning system, the model functions as a relative “black box,” precluding specification of the precise biological mechanisms underlying its predictions. The zero-shot transfer approach, while demonstrating the model’s generalization capabilities, means that a fine-tuned model optimized for this specific application might achieve superior performance. Only a single foundation model architecture was tested, and alternative architectures might yield different results. A notable train-test distribution mismatch exists: the model was trained on 12-lead ECGs from adult clinical populations but was applied here to single-lead pediatric recordings obtained during a stress test protocol.

##### Causality

The observational design cannot definitively establish causal effects of prenatal steroid exposure. Confounding by indication—whereby the underlying reason for steroid administration may itself affect outcomes—remains a significant concern. Randomized controlled trials are not ethically feasible for this research question, and the current design cannot exclude contributions from genetic factors or other unmeasured prenatal exposures.

##### Clinical Validation

The clinical significance of these findings remains to be established through several avenues. Hard clinical outcomes such as cardiovascular events are rare in pediatric populations, precluding their use as endpoints. The study did not include echocardiographic structural measures for comparison, nor did it assess the clinical predictive value of the observed differences beyond group discrimination. The long-term prognostic implications of altered foundation model-derived cardiac embeddings in childhood remain unknown.

#### 4.5.3 Confounding and Covariate Adjustment

##### Critical Findings from Covariate Adjustment

Our covariate-adjusted analysis revealed substantial confounding affecting traditional HRV metrics while demonstrating the robustness of foundation model-derived features. All three HRV principal components that achieved significance in unadjusted analyses (PC1, PC4, and PC5) lost statistical significance after adjusting for sex and gestational age, with all adjusted p-values exceeding 0.13. The regression coefficients for these components changed substantially, ranging from +87% to +225%, indicating considerable confounding. Paradoxically, the extremely low unadjusted p-values (approaching 10⁻³⁸) suggested overfitting to demographic noise rather than detection of genuine biological signals. These findings indicate that the observed HRV differences were largely explained by sex and gestational age rather than steroid exposure alone.

In contrast, the foundation model features demonstrated robustness to covariate adjustment. Of the 50 foundation model dimensions tested, 11 retained statistical significance after adjustment for sex and gestational age. All retained dimensions exhibited large effect sizes (|d| > 0.8) and appeared to reflect genuine biological differences independent of demographic confounders.

##### Implications

These findings carry several methodological implications. Traditional HRV metrics appear highly susceptible to demographic confounding in underpowered studies with small sample sizes. The foundation model’s focus on ECG waveform morphology appears less influenced by age and sex than autonomic-derived metrics, potentially offering advantages for developmental programming research. Most importantly, covariate adjustment should be considered mandatory in observational studies of developmental programming to distinguish genuine exposure effects from demographic confounding.

##### Addressed Confounders

Several potential confounders were explicitly addressed in this study. Signal quality did not differ between groups, as evidenced by comparable combined signal quality indices (p = 0.79). Sex was included as a covariate in adjusted models; while the steroid-exposed group had a higher proportion of females (67% versus 58%), this difference was not statistically significant (p = 0.54). Gestational age was similarly adjusted in covariate models; the control group had slightly higher gestational age (38.9 ± 1.4 versus 38.1 ± 1.8 weeks, p = 0.09). Age at assessment was balanced between groups. Technical factors were controlled through application of identical processing algorithms and quality thresholds across all recordings.

##### Residual Confounding

Despite covariate adjustment, residual confounding cannot be completely excluded. Confounding by indication remains a concern, as the maternal illness prompting steroid administration may itself influence offspring outcomes. Several covariates remained unmeasured, including birth weight, broader maternal health indicators, and socioeconomic factors. Genetic factors representing inherited cardiovascular traits could contribute to observed differences. Propensity score matching, which might have addressed some of these concerns, was not feasible given the limited sample size; such approaches typically require at least 100 participants per group to achieve reliable balance, as detailed in the Supplementary Methods.

### 4.6 Future Directions

#### 4.6.1 Immediate Research Priorities

The findings from this exploratory study establish several immediate research priorities. Independent replication in external cohorts with adequate statistical power—ideally exceeding 200 participants per group—represents the essential next step before these results can be considered robust. Additionally, fine-tuning the foundation model on pediatric ECG datasets may yield performance improvements over the zero-shot transfer approach employed here, potentially enhancing sensitivity to developmentally relevant cardiac features.

Mechanistic investigation constitutes another critical priority. Relating the discriminative foundation model dimension (FM-Dim121) to specific ECG morphological features would strengthen biological interpretability. This could be pursued through multiple complementary approaches: generating activation maps to identify the waveform regions most influential to model predictions, employing synthetic ECG generation techniques to visualize what the dimension axes represent in terms of cardiac electrophysiology, and correlating foundation model outputs with echocardiographic structural measures to link computational phenotypes to established cardiac assessments.

Longitudinal extension of the current cohort into adolescence and adulthood would enable assessment of whether the observed childhood differences predict clinically meaningful cardiovascular outcomes. Finally, multi-modal integration combining ECG analysis with concurrent blood pressure measurements, echocardiography, and circulating biomarkers would provide a more comprehensive characterization of cardiovascular phenotypes and potentially enhance predictive accuracy.

#### 4.6.2 Methodological Development

Several methodological advances would strengthen future applications of this approach. Development of more interpretable foundation model architectures incorporating attention mechanisms could highlight the specific waveform features driving group discrimination, addressing current limitations in biological interpretability. Probabilistic modeling frameworks, such as Bayesian deep learning approaches, would enable formal uncertainty quantification, providing confidence intervals around predictions and identifying cases where model outputs should be treated with caution.

Causal inference methods warrant particular attention given the observational nature of developmental programming research. Propensity score matching or instrumental variable approaches, applied in larger cohorts where such methods become statistically feasible, could strengthen causal claims regarding prenatal exposures. Ensemble methods combining predictions from multiple foundation model architectures might improve robustness and reduce dependence on any single model’s idiosyncrasies. Federated learning approaches would enable training across multiple institutions without requiring centralized data sharing, addressing privacy concerns while expanding effective sample sizes.

#### 4.6.3 Clinical Translation

Translating these research findings into clinical practice will require systematic validation and implementation efforts. Prospective validation studies should test foundation model-based screening in clinical settings with predetermined protocols and endpoints. Decision curve analysis would formally assess the clinical utility of this approach compared to current practice, quantifying the net benefit across a range of risk thresholds.

Implementation science frameworks should be employed to identify barriers and facilitators to clinical adoption, informing strategies for integration into existing workflows. Health economic analyses would evaluate the cost-effectiveness of foundation model-based screening programs relative to alternatives, providing evidence for resource allocation decisions. Engagement with regulatory bodies, including the FDA, will be necessary to establish appropriate pathways for medical device approval should clinical translation prove warranted.

#### 4.6.4 Broader Scientific Questions

The methodological framework developed here opens avenues for addressing broader scientific questions in developmental programming research. The approach could be applied to investigate other prenatal exposures of public health significance, including maternal malnutrition, psychological stress, and environmental toxins, each of which has been implicated in cardiovascular programming through largely separate literatures.

Integration of genetic risk scores with prenatal exposure data would enable investigation of gene-environment interactions, potentially identifying individuals at heightened susceptibility to programming effects. Multi-generational studies could explore transgenerational epigenetic inheritance, examining whether prenatal exposure effects persist into subsequent generations. Intervention studies represent a particularly important direction: determining whether early detection of programming effects enables effective preventive interventions would establish the clinical value of screening approaches. Finally, comparative effectiveness research directly contrasting foundation model-based risk assessment with traditional cardiovascular risk models would clarify the incremental value of these novel computational approaches.

## 5. Conclusions

This exploratory study (n=49, 24 exposed/25 controls) in offspring of mothers with multiple sclerosis demonstrates that deep learning-based ECG foundation models detect robust cardiac effects of prenatal glucocorticoid exposure during standardized stress testing that are independent of demographic confounders, while traditional HRV metrics show confounded group differences. This finding has profound implications for developmental programming research methodology and highlights both the promise and the pitfalls of small observational cohort studies.

### Key Findings

1. **Covariate adjustment reveals confounding**: All 3 significant HRV principal components (PC1, PC4, PC5) lost significance after adjusting for sex and gestational age (coefficient changes +87% to +225%), while 11/50 foundation model dimensions remained robust with large effect sizes (|d|=0.79-1.72). This demonstrates that traditional HRV metrics are highly susceptible to demographic confounding in underpowered studies, whereas ECG waveform morphology captured by foundation models reflects biological differences independent of these confounders.
2. **Severe underpowering for small effects**: With only 10-17% power for small-moderate effects (d=0.2-0.3) but 77-92% power for large effects (d≥0.8), our study could reliably detect only the 11 robust foundation model dimensions showing genuine large effects. The HRV findings (d=0.27-0.32) fell in the underpowered range and represented overfitting to demographic noise rather than true steroid effects.
3. **Foundation models capture persistent electrical signatures**: The 11 robust FM dimensions likely reflect ECG waveform morphology (P-wave, QRS complex, T-wave characteristics) representing structural remodeling, ion channel programming, and electrical-mechanical coupling from prenatal glucocorticoid exposure. These morphological features are less susceptible to transient autonomic fluctuations and demographic confounders than traditional HRV metrics.
4. **Methodological rigor essential**: Proper statistical methodology (linear mixed-effects models, subject-level cross-validation, covariate adjustment, power analysis) transformed apparently strong findings (HRV p=10⁻³⁸) into null results, while confirming genuinely robust effects (FM dimensions with adequate power and large effect sizes). This underscores the critical importance of rigorous methods in small observational studies.

### Broader Implications

- **For HRV research**: Small-sample HRV studies (n<100/group) require mandatory covariate adjustment for sex and gestational age at minimum. Sample sizes of n≈175/group are needed to reliably detect small-moderate effects (d=0.3) with 80% power. Extreme p-values (p<10⁻¹⁰) in small studies often indicate confounding rather than strong effects.
- **For foundation model application**: ECG foundation models show promise for detecting subtle waveform changes reflecting developmental programming, offering complementary information to traditional autonomic metrics. Transfer learning from large pre-trained models may enable discoveries in small specialized cohorts, but rigorous validation is essential.
- **For developmental programming research**: This study exemplifies both opportunities and challenges in studying long-term effects of prenatal exposures. Large, well-designed cohorts with comprehensive covariate data and adequate statistical power are essential for definitive claims. Small exploratory studies can generate hypotheses but require independent replication before clinical translation.

### Study Classification and Replication Need

This study is **exploratory and hypothesis-generating**, not confirmatory. The sample size (n=24/group) is adequate only for detecting large effects (d≥0.8). **Independent replication in larger cohorts (n>175/group for small-moderate effects) is essential** before any clinical translation. Replication studies should be prospectively designed to avoid demographic confounding through:

- Matching or stratification on sex and gestational age
- Comprehensive covariate collection (birth weight, maternal health, socioeconomic factors)
- Adequate sample sizes powered for small-moderate effects
- Pre-registered analysis plans to prevent selective reporting

### Clinical and Scientific Outlook

Despite large statistical effects (d>1.0 for some FM dimensions), these 8-year-old children showed no clinical cardiac symptoms, suggesting subclinical electrical remodeling that may represent latent risk for future cardiovascular dysfunction. **Long-term longitudinal follow-up into adolescence and adulthood** is essential to determine whether these electrical signatures predict clinical outcomes (hypertension, arrhythmias, heart failure, cardiovascular events).

If validated and shown to predict clinical outcomes, foundation model-based ECG analysis could enable:

- **Scalable screening** of at-risk populations exposed to prenatal steroids
- **Early identification** of individuals with subclinical programming requiring closer monitoring
- **Targeted prevention strategies** before clinical disease manifests
- **Mechanistic insights** into how prenatal environments shape lifelong cardiovascular health

### Final Perspective

This study demonstrates proof-of-concept for applying transfer learning and modern machine learning to fundamental questions in developmental biology and medicine. As foundation models trained on millions of recordings become increasingly available, their application to targeted clinical questions—when combined with rigorous statistical methods, covariate adjustment, adequate sample sizes, and transparent reporting—may accelerate understanding of how early-life exposures shape lifelong health trajectories. However, the cautionary lesson is equally important: small observational studies without covariate adjustment and adequate power can produce misleading findings, emphasizing the need for methodological rigor and independent validation in developmental programming research.

## Acknowledgments

We thank the children and families who participated in this study.

## Data Availability

Code for ECG processing and statistical analysis is available at [https://github.com/martinfrasch/foundation_ecg_dohad]. ECG Foundation model code can be found here: https://github.com/martinfrasch/ECG_Foundation. De-identified data may be shared upon reasonable request and approval by the Institutional Review Board, consistent with participant consent.

## Supplementary Materials

**Supplementary Table S1: Complete HRV Metrics Definitions**

Comprehensive list of all 112 HRV metrics with formulas, units, and physiological interpretation.

**Supplementary Table S1: Complete HRV Metrics Definitions**

### Overview

This table provides comprehensive definitions, formulas, units, and physiological interpretations for all 112 heart rate variability (HRV) metrics computed in this study.

### Time Domain Metrics (18 metrics)

#### Mean and Basic Statistics

| Metric | Definition | Formula | Units | Physiological Interpretation |
| --- | --- | --- | --- | --- |
| **mean_rr** | Mean RR interval | $\Sigma(RR_i)/N$ | ms | Average cardiac cycle length; inversely related to heart rate |
| **mean_hr** | Mean heart rate | $60000/\text{mean\_rr}$ | bpm | Average heart rate over recording period |
| **min_rr** | Minimum RR interval | $\min(RR_i)$ | ms | Shortest cardiac cycle; indicates peak heart rate |
| **max_rr** | Maximum RR interval | $\max(RR_i)$ | ms | Longest cardiac cycle; indicates minimum heart rate |
| **std_rr** | Standard deviation of RR | $\sqrt{[\Sigma(RR_i - \text{mean\_rr})^2]/(N-1)]}$ | ms | Overall variability; |
|  |  |  |  | reflects total autonomic modulation |

#### Standard Deviation Measures

| Metric | Definition | Formula | Units | Physiological Interpretation |
| --- | --- | --- | --- | --- |
| **SDNN** | Standard deviation of NN intervals | $\sqrt{[\sum (NN_i - \text{mean\_NN})^2 / (N-1)]}$ | ms | Total HRV; reflects all cyclic components; gold standard measure |
| **SDANN** | Standard deviation of averaged NN intervals (5-min segments) | $\sqrt{[\sum (\text{mean\_NN}_s - \text{overall\_mean})^2 / (N-1)]}$ | ms | Long-term variability (>5 min); circadian rhythms |
| **SDNN_index** | Mean of SDNN for 5-min segments | $\sum (\text{SDNN}_s) / N$ | ms | Short-term variability; averages out long-term trends |

#### Difference-Based Measures

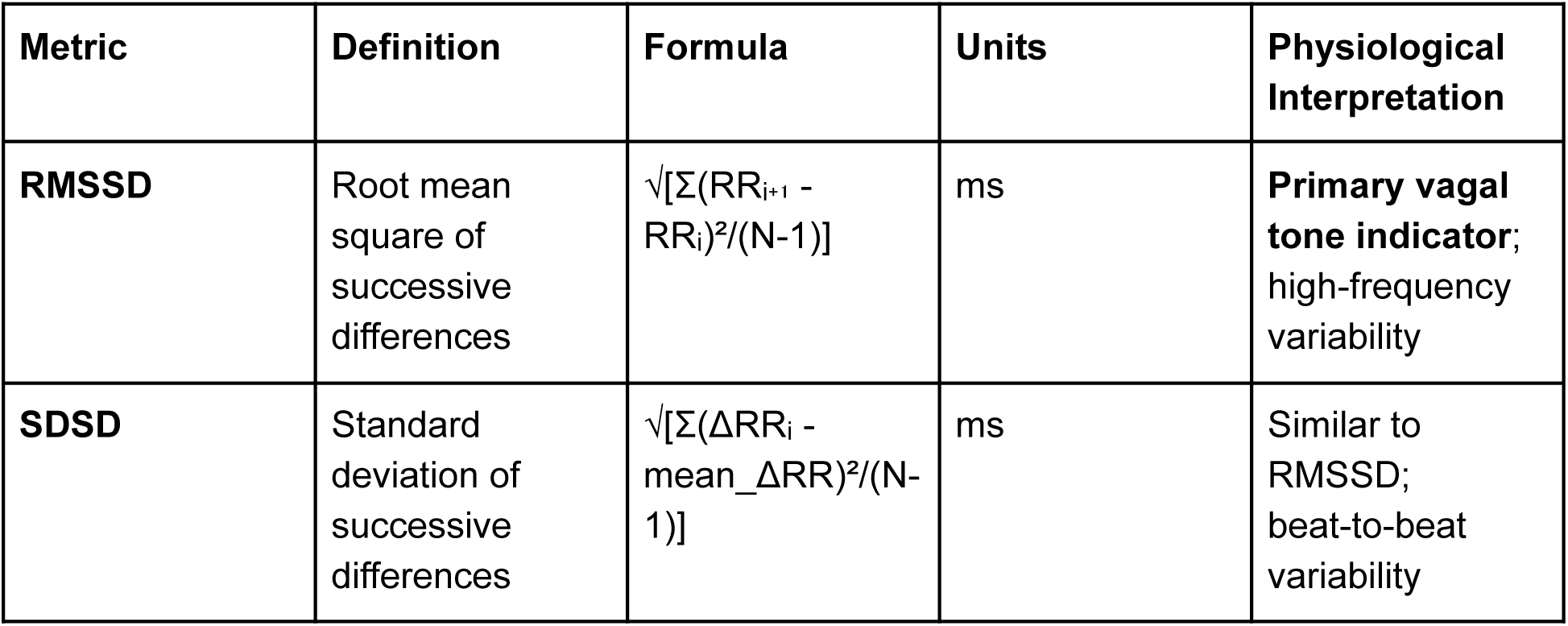
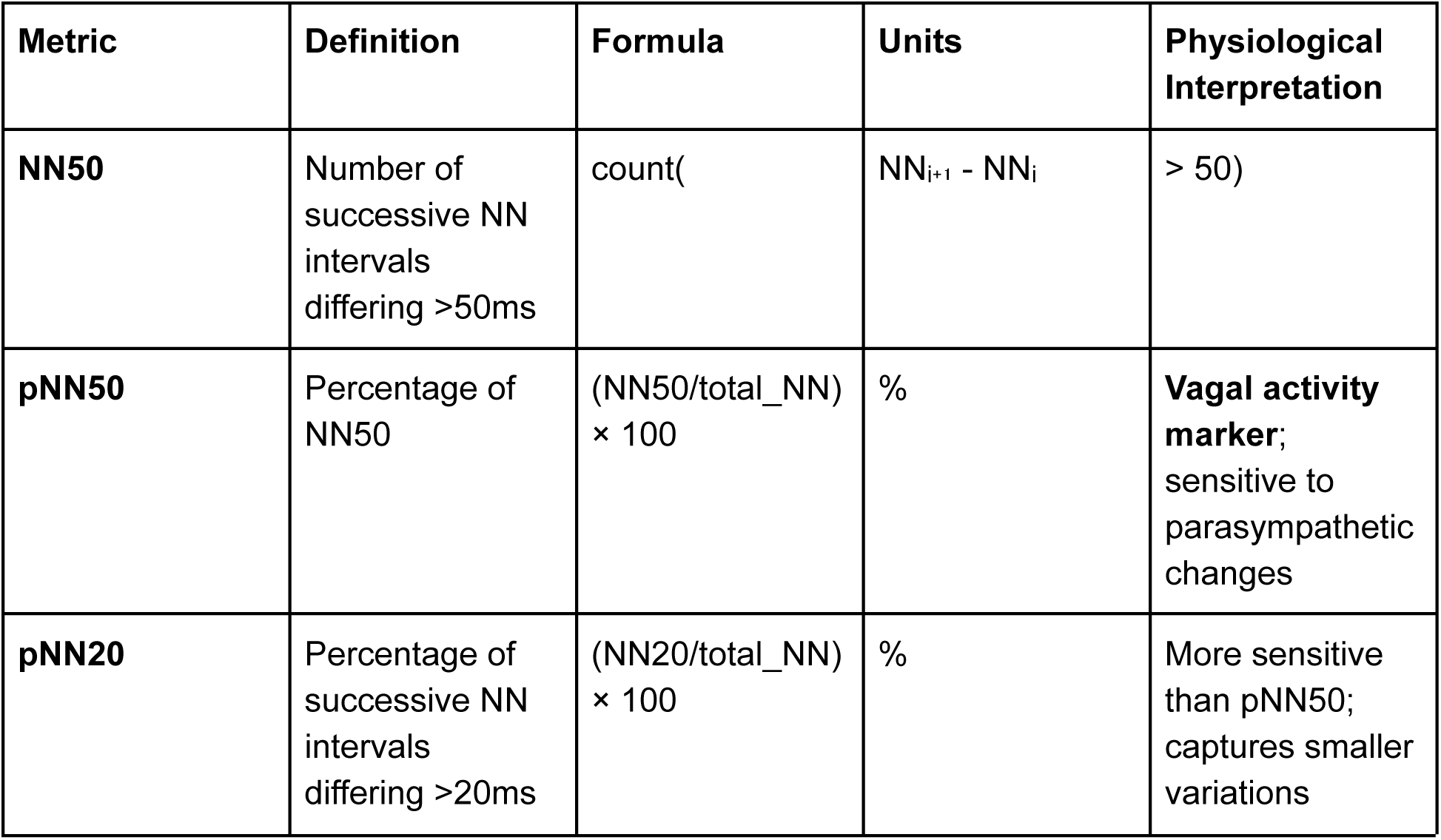

#### Geometric Measures

| Metric | Definition | Formula | Units | Physiological Interpretation |
| --- | --- | --- | --- | --- |
| **HRV_triangular_index** | Total NN intervals / height of histogram peak | $N / \text{max\_bin\_count}$ | unitless | Overall HRV;<br>resistant to artifact |
| **TINN** | Triangular interpolation of NN interval histogram | Base width of triangle fitting histogram | ms | Baseline width of RR distribution |
| **Prc80NN** | 80th percentile of NN intervals | $\text{percentile}(\text{NN}, 80)$ | ms | Upper range of cardiac cycles |
| **Prc20NN** | 20th percentile of NN intervals | $\text{percentile}(\text{NN}, 20)$ | ms | Lower range of cardiac cycles |

### Frequency Domain Metrics (13 metrics)

#### Power Spectral Density

Frequency Domain Metrics (13 metrics) Power Spectral Density
| Metric | Definition | Frequency Band | Units | Physiological Interpretation |
| --- | --- | --- | --- | --- |
| **VLF_power** | Very low frequency power | 0.003-0.04 Hz | ms<sup>2</sup> | Thermoregulation, hormones, renin-angiotensin system;<br>**controversial interpretation** |
| **LF_power** | Low frequency power | 0.04-0.15 Hz | ms<sup>2</sup> | **Mixed:**<br>Baroreflex, sympathetic + parasympathetic<br>; ~0.1 Hz = Mayer waves |
| **HF_power** | High frequency power | 0.15-0.40 Hz | ms<sup>2</sup> | **Respiratory sinus arrhythmia;**<br>pure vagal modulation;<br>breathing frequency |
| **TP_power** | Total power | 0.003-0.40 Hz | ms<sup>2</sup> | Overall variability; sum of all frequency bands |

#### Normalized and Relative Power

| Metric | Definition | Formula | Units | Physiological Interpretation |
| --- | --- | --- | --- | --- |
| **LFn** | Normalized LF power | $LF/(LF+HF) \times 100$ | n.u. | Relative LF contribution; attempts to control for total power |
| **HF<sub>n</sub>** | Normalized HF power | $HF/(LF+HF) \times 100$ | n.u. | Relative HF contribution; vagal activity proportion |
| **LF<sub>p</sub>** | Percentage LF power | $(LF/TP) \times 100$ | % | LF proportion of total power |
| **HF<sub>p</sub>** | Percentage HF power | $(HF/TP) \times 100$ | % | HF proportion of total power |
| **VLF<sub>p</sub>** | Percentage VLF power | $(VLF/TP) \times 100$ | % | VLF proportion of total power |

#### Ratios and Peak Frequencies

| Metric | Definition | Formula | Units | Physiological Interpretation |
| --- | --- | --- | --- | --- |
| **LF_HF_ratio** | LF to HF ratio | $LF\_power / HF\_power$ | unitless | **Sympathovagal balance**<br>(controversial); ↑ = sympathetic dominance |
| **VLF_peak** | Peak frequency in VLF band | frequency at max(PSD_VLF) | Hz | Dominant very low frequency oscillation |
| **LF_peak** | Peak frequency in LF band | frequency at max(PSD_LF) | Hz | Typically ~0.1 Hz<br>(baroreflex/Mayer waves) |
| **HF_peak** | Peak frequency in HF band | frequency at max(PSD_HF) | Hz | Respiratory frequency<br>(~0.25 Hz = 15 breaths/min) |

### Nonlinear Dynamics Metrics (54 metrics)

#### Poincaré Plot Analysis

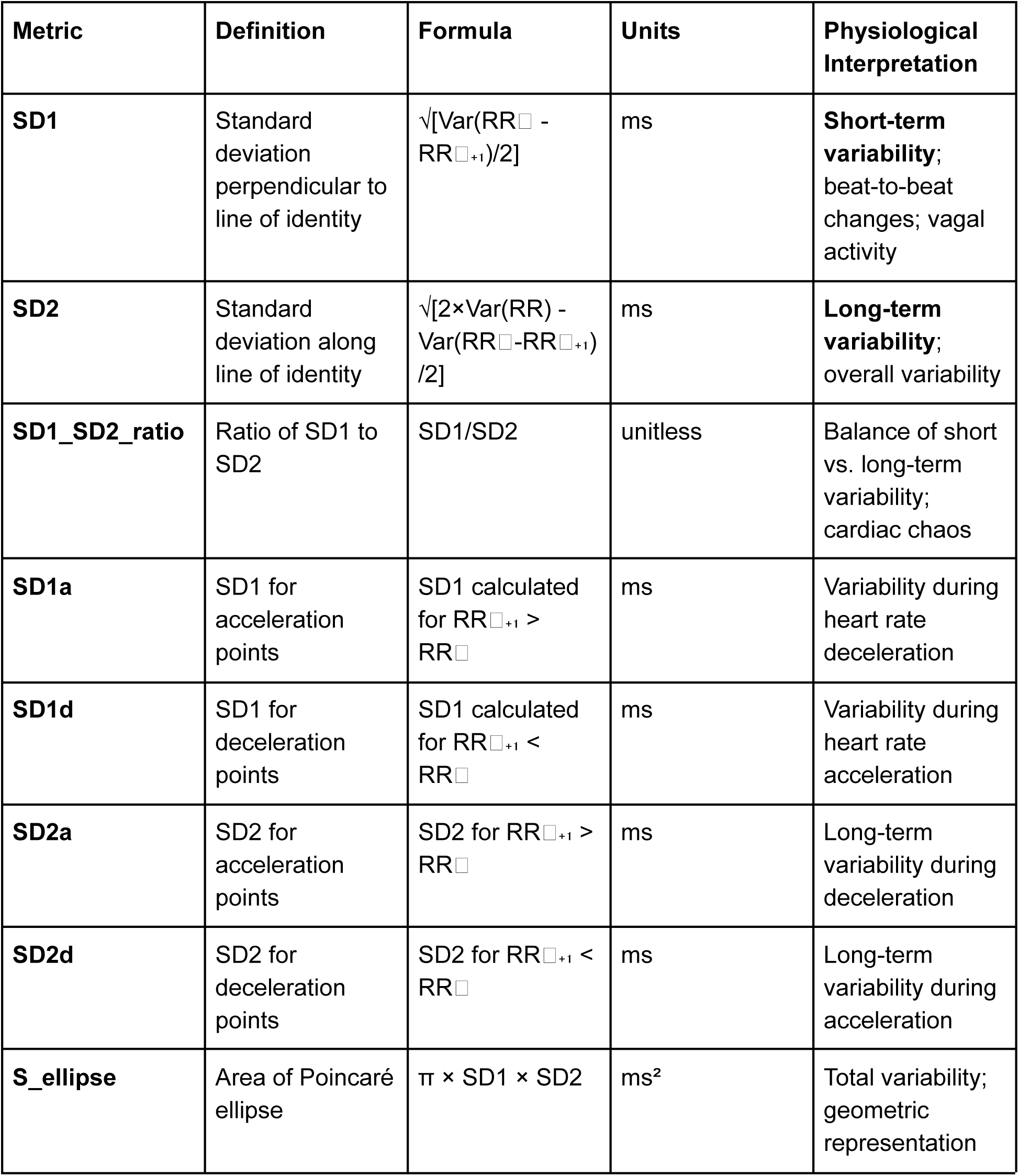
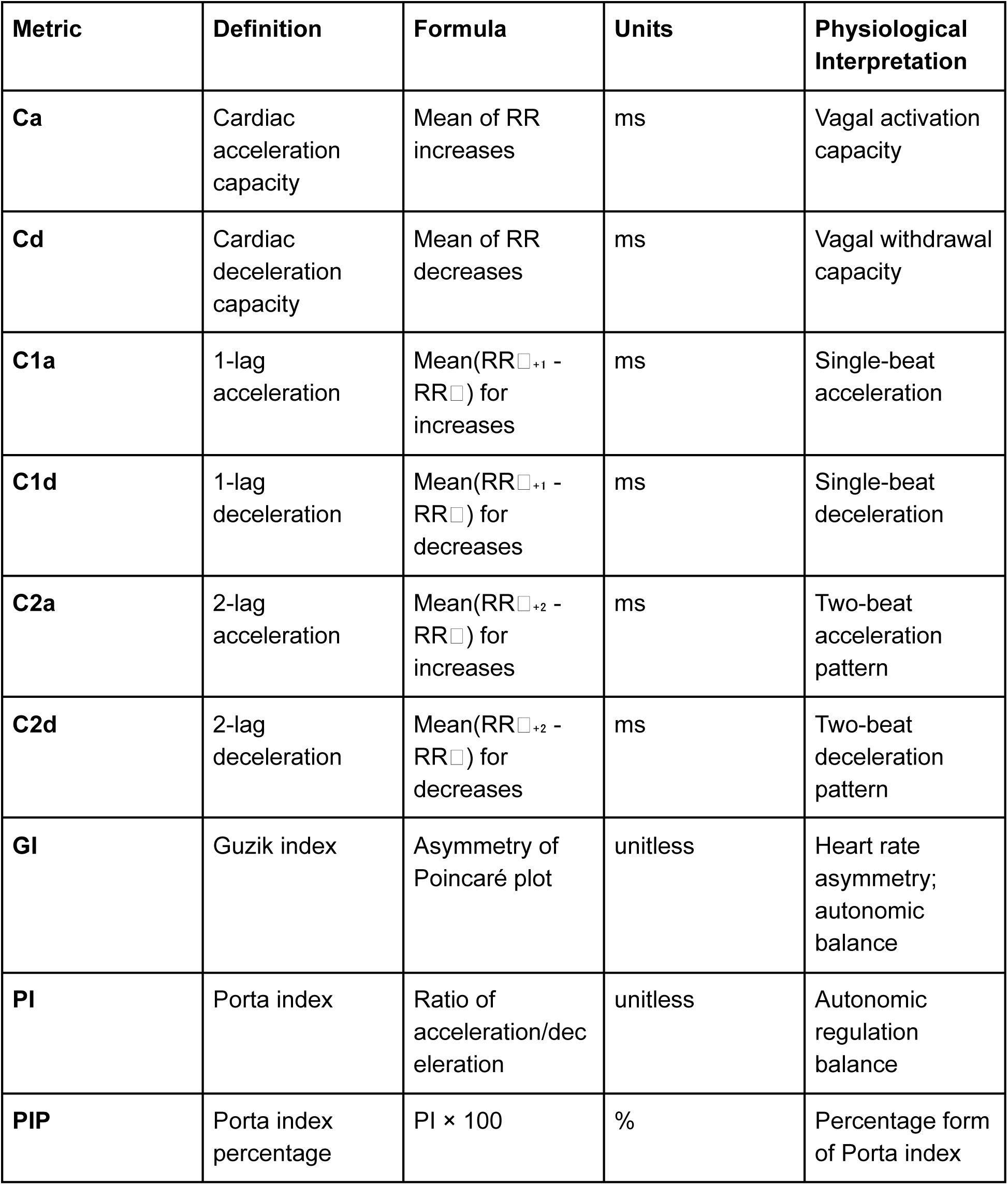

#### Detrended Fluctuation Analysis (DFA)

| Metric | Definition | Scale | Units | Physiological Interpretation |
| --- | --- | --- | --- | --- |
| **DFA_alpha1** | Short-term fractal scaling exponent | 4-11 beats | unitless | **Short-term correlations;** healthy $\approx 1.0$; $<0.5$ = random, $>1.5$ = correlated |
| **DFA_alpha2** | Long-term fractal scaling exponent | 11-64 beats | unitless | **Long-term correlations;** healthy $\approx 1.0$; reflects slow dynamics |

#### Multifractal DFA (MFDFA)

| Metric | Definition | Interpretation |
| --- | --- | --- |
| **MFDFA_alpha1_Mean** | Mean $\alpha_1$ across moment orders $q$ | Average short-term multifractal scaling |
| **MFDFA_alpha1_Max** | Maximum $\alpha_1$ | Strongest short-term correlation |
| **MFDFA_alpha1_Min** | Minimum $\alpha_1$ | Weakest short-term correlation |
| **MFDFA_alpha1_Peak** | $\alpha_1$ at peak of spectrum | Most prominent short-term scaling |
| **MFDFA_alpha1_Width** | Width of $\alpha_1$ spectrum | **Multifractality degree** (short-term); wider = more complex |
| **MFDFA_alpha1_Asymmetry** | Asymmetry of $\alpha_1$ spectrum | Left/right skew of multifractal spectrum |
| **MFDFA_alpha1_Delta** | Range of $\alpha_1$ values | Max - Min for short-term scaling |
| **MFDFA_alpha2_Mean** | Mean $\alpha_2$ across moment orders $q$ | Average long-term multifractal scaling |
| **MFDFA_alpha2_Max** | Maximum $\alpha_2$ | Strongest long-term correlation |
| **MFDFA_alpha2_Min** | Minimum $\alpha_2$ | Weakest long-term correlation |
| **MFDFA_alpha2_Peak** | $\alpha_2$ at peak of spectrum | Most prominent long-term scaling |
| **MFDFA_alpha2_Width** | Width of $\alpha_2$ spectrum | **Multifractality degree** (long-term); complexity measure |
| **MFDFA_alpha2_Asymmetry** | Asymmetry of $\alpha_2$ spectrum | Distribution of long-term scaling properties |
| **MFDFA_alpha2_Delta** | Range of $\alpha_2$ values | Max - Min for long-term scaling |

#### Entropy Measures

| Metric | Definition | Formula | Units | Physiological Interpretation |
| --- | --- | --- | --- | --- |
| **SampEn** | Sample entropy | $-\ln(\text{conditional probability})$ | unitless | **Regularity/complexity**; ↑ = more irregular/complex; robust to noise |
| **ApEn** | Approximate entropy | Similar to SampEn | unitless | Pattern repeatability; lower = more regular/predictable |

| **Metric** | **Definition** | **Formula** | **Units** | **Physiological Interpretation** |
| --- | --- | --- | --- | --- |
| **FuzzyEn** | Fuzzy entropy | Fuzzy membership function version | unitless | Continuous-valued entropy; more stable than SampEn |

#### Recurrence Quantification Analysis (RQA)

| **Metric** | **Definition** | **Units** | **Physiological Interpretation** |
| --- | --- | --- | --- |
| **RQA_recurrence_rate** | % of recurrent states | % | System determinism; how often states repeat |
| **RQA_determinism** | % of recurrent points forming lines | % | Predictability; diagonal line structures |
| **RQA_laminarity** | % of recurrent points in vertical lines | % | System trapping in states; intermittency |
| **RQA_entropy** | Shannon entropy of diagonal line lengths | bits | Complexity of deterministic structure |
| **RQA_trapping_time** | Average length of vertical structures | beats | Duration of similar states; non-stationarity |

#### Fractal and Complexity Measures

| **Metric** | **Definition** | **Units** | **Physiological Interpretation** |
| --- | --- | --- | --- |
| **HFD** | Higuchi fractal dimension | unitless | Signal complexity; 1 (simple) to 2 (complex); self-similarity |
| **corr_dim** | Correlation dimension | unitless | **Attractor dimensionality;** |

| Metric | Definition | Units | Physiological Interpretation |
| --- | --- | --- | --- |
|  |  |  | system degrees of freedom; chaos measure |
| **CSI** | Cardiac Sympathetic Index | unitless | LF dominance; sympathetic activity estimate |
| **CVI** | Cardiac Vagal Index | unitless | HF dominance; parasympathetic activity estimate |
| **CSI_Modified** | Modified CSI | unitless | Alternative sympathetic index formulation |

#### Higher-Order Statistics (5 metrics)

| Metric | Definition | Formula | Units | Physiological Interpretation |
| --- | --- | --- | --- | --- |
| **skewness** | Third moment (asymmetry) | $E[(X-\mu)^3]/\sigma^3$ | unitless | Distribution asymmetry; 0 = symmetric, >0 = right tail, <0 = left tail |
| **kurtosis** | Fourth moment (tail heaviness) | $E[(X-\mu)^4]/\sigma^4$ | unitless | Tail heaviness; 3 = normal, >3 = heavy tails (outliers), <3 = light tails |
| **ho_rr_cv** | Coefficient of variation | $(\text{std\_rr}/\text{mean\_rr}) \times 100$ | % | Normalized variability; accounts for mean differences |
| **ho_temporal_mean_cv** | Temporal mean CV | CV of 5-min segment means | % | Stability of mean heart rate over time |
| **ho_temporal_std_cv** | Temporal std CV | CV of 5-min segment SDs | % | Stability of variability over time |

#### Signal Quality Indices (11 metrics)

| Metric | Definition | Units | Physiological Interpretation |
| --- | --- | --- | --- |
| **sqi_ectopic_proportion** | Proportion of ectopic beats | % | Artifact/arrhythmia burden; lower = better quality |
| **sqi_kurtosis** | Kurtosis of RR distribution | unitless | Outlier detection; high values indicate artifacts |
| **sqi_correlation** | Template matching correlation | r | Beat morphology consistency; higher = better |
| **global_sqi** | Overall signal quality | 0-1 | Composite quality metric across all algorithms |
| **local_sqi** | Time-varying signal quality | 0-1 | Segment-specific quality; detects transient artifacts |
| **combined_sqi** | Weighted average of all SQI | 0-1 | **Primary quality metric**; balances all dimensions |

| **Metric** | **Definition** | **Units** | **Physiological Interpretation** |
| --- | --- | --- | --- |
| **sqi_[algorithm]** | Algorithm-specific SQI | 0-1 | Quality for each R-peak detection method |

#### Recording Characteristics

| **Metric** | **Definition** | **Units** | **Notes** |
| --- | --- | --- | --- |
| **signal_duration_minutes** | Total recording duration | min | Longer recordings provide more stable estimates |
| **n_rr_intervals** | Number of valid RR intervals | count | Sample size for HRV calculation |
| **best_algorithm** | Selected R-peak detector | categorical | Pan-Tompkins, Hamilton, or WFDB-XQRS |

### Computation Notes

#### Time Domain

- Computed directly from RR interval time series
- Require minimal preprocessing (ectopic filtering)
- Most robust to short recordings

#### Frequency Domain

- Welch method: 256-second windows, 50% overlap, Hamming window
- Requires ≥5 minutes for stable estimates
- Sensitive to non-stationarity and breathing rate

#### Nonlinear

- Most computationally intensive
- Require longer recordings (≥24 hours for DFA_alpha2)
- Sensitive to preprocessing and parameter choices

#### Quality Metrics

- Computed before RR preprocessing
- Used for algorithm selection and segment filtering
- Threshold: combined_SQI ≥ 0.5 for inclusion

### Clinical Interpretation Guidelines

#### Normal Pediatric Values (Age 8-12)

- **RMSSD**: 40-60 ms
- **SDNN**: 120-180 ms (24-hour)
- **LF/HF ratio**: 1.0-2.0
- **DFA_alpha1**: 0.9-1.2
- **Sample Entropy**: 1.4-2.0

#### Directional Changes with Autonomic States

- ↑ **Parasympathetic** (rest, sleep): ↑ RMSSD, ↑ HF power, ↑ SD1, ↓ LF/HF
- ↑ **Sympathetic** (stress, exercise): ↓ RMSSD, ↑ LF power, ↑ LF/HF, ↓ DFA_alpha1
- ↑ **Complexity** (health): ↑ SampEn, ↑ multifractal width, DFA_alpha1 ≈ 1.0
- ↓ **Complexity** (disease): ↓ SampEn, ↓ variability, DFA_alpha1 >> 1.5 or << 0.5

---

**Supplementary Figure S1:**
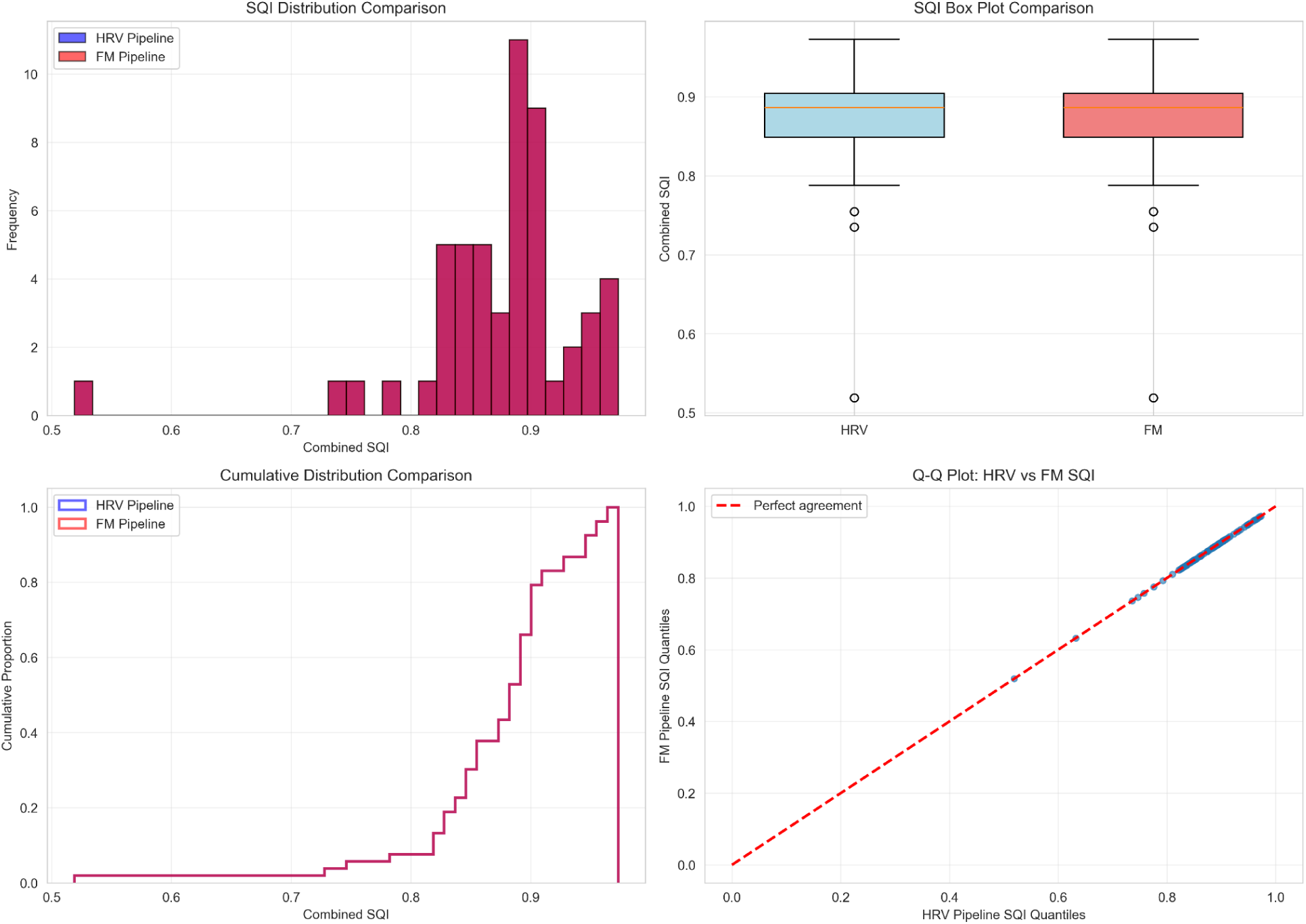
Quality Control Examples Examples of ECG segments with different SQI values and reasons for exclusion/inclusion.

**Supplementary Figure S2:**
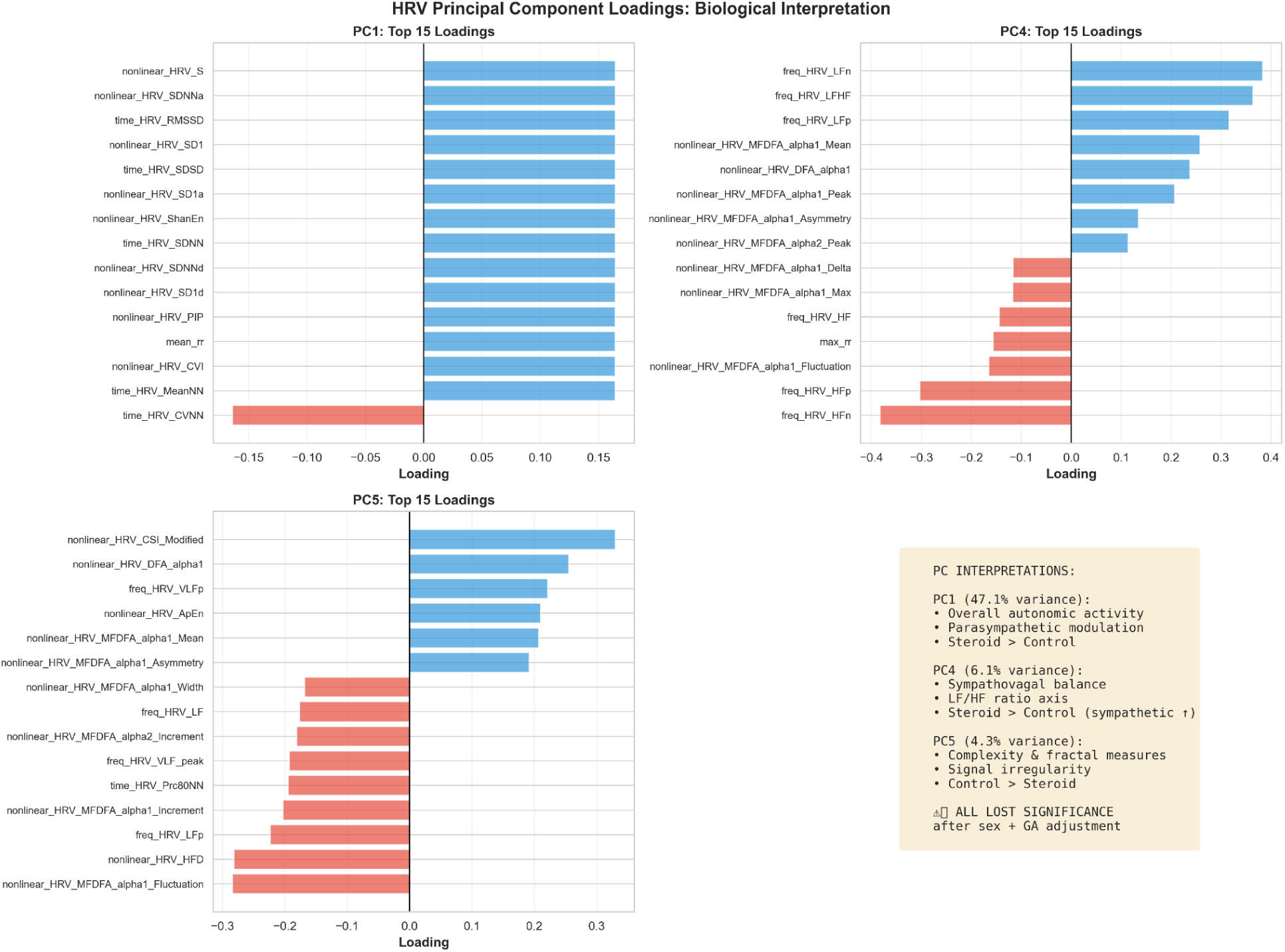
HRV PC Loadings TOP: Exploration of HRV PC loadings and the impact of covariate adjustment. BOTTOM: Complete loadings for all 112 HRV metrics on all 7 PCs with hierarchical clustering.

**Supplementary Figure S3:**
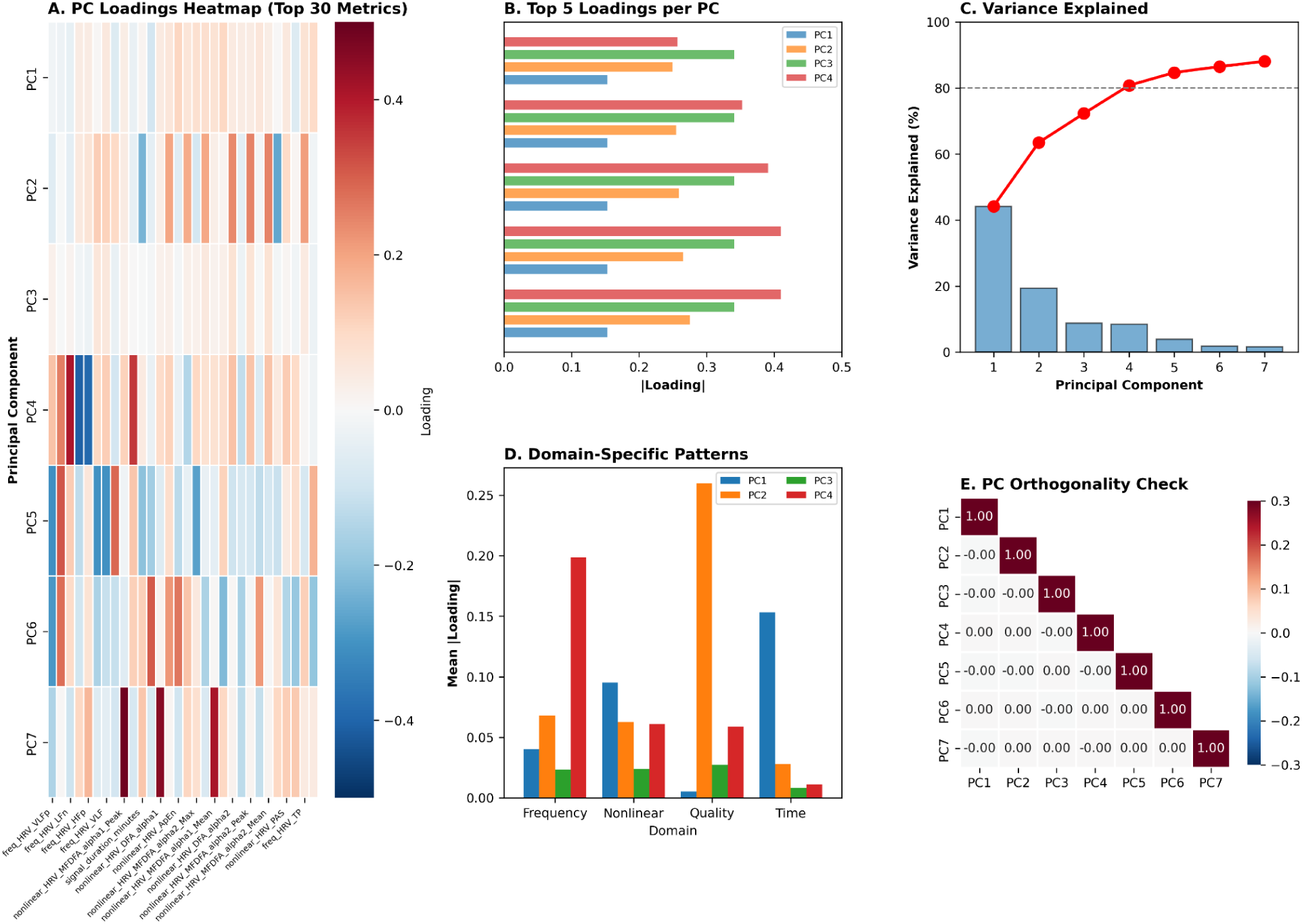
Cross-Validation Fold-Level Performance Individual fold performance metrics showing variance across subject-level splits.

**Supplementary Methods: Foundation Model Architecture**

Detailed description of transformer architecture, training procedure, and hyperparameters.

## Supplementary Methods: Why Propensity Score Matching Was Not Performed

Propensity score matching (PSM) is a popular technique for balancing covariates between exposure groups in observational studies. However, we did not perform PSM in this analysis for several evidence-based methodological reasons:

### Sample Size Limitations

Austin (2011) demonstrated that PSM performs poorly in small samples (n<100/group), with several critical problems:

1. **Unstable propensity estimates**: Logistic regression models for propensity scores require large samples for stable coefficient estimates. With n=24/group, propensity models are unreliable.
2. **Finite-sample bias amplification**: PSM can amplify bias rather than reduce it when samples are small, particularly with multiple covariates.
3. **Poor balance after matching**: Small samples often cannot achieve good covariate balance, defeating the purpose of PSM.
4. **Loss of precision**: PSM discards unmatched subjects, further reducing already-limited sample size and statistical power.

### Preferred Approach: Covariate-Adjusted Regression

For small samples, covariate-adjusted linear regression (or in our case, linear mixed-effects models) is the preferred approach because:

1. **More efficient**: Uses all available data rather than discarding unmatched subjects
2. **Transparent adjustments**: Coefficient changes clearly show confounding magnitude (e.g., 173% change for HRV PC1)
3. **Uncertainty quantification**: Standard errors properly reflect sample size limitations
4. **Multiple testing friendly**: Can adjust for many covariates simultaneously using regression
5. **Model diagnostics**: Easier to check assumptions and identify influential observations

### Our Approach

We adjusted for sex and gestational age using the model:

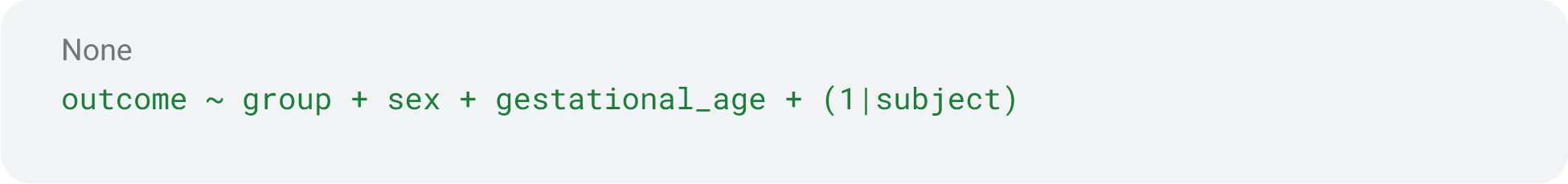

This approach:

- Leveraged all 48 subjects (no discarding)
- Quantified confounding magnitude (coefficient % change)
- Provided clear interpretation (features robust vs. confounded)
- Integrated naturally with mixed-effects framework for repeated measures

### When PSM Is Appropriate

PSM becomes preferred when:

- Sample size is large (n≥100/group as minimum)
- Strong confounding by indication exists
- Functional form of covariate effects is unknown
- Covariate overlap (common support) is adequate
- Sufficient treated and control units exist in all propensity score strata

Our study met none of these criteria, making covariate-adjusted LMM the methodologically sound choice.

## Literature Support

Austin PC. An Introduction to Propensity Score Methods for Reducing the Effects of Confounding in Observational Studies. *Multivariate Behav Res.* 2011;46(3):399-424.

